# A world model simulates the latent dynamics of human health

**DOI:** 10.64898/2026.09.19.26363460

**Authors:** Zirui Wang, Odin Zhang, Jiaqi Wang, Yijia Jiang, Zijian Carl Ma, Minghao Guo, Saleem Al Dajani, Alec Eames, Alibek Moldakozhayev, Jesse R. Poganik, Mahdi Moqri, Dane Gobel, Ranran Zhai, Alina Isakova, Yingcheng Wu, Zhenfei Yin, Le Cong, George Slavich, Wojciech Matusik, Hannele Ruohola-Baker, James Zou, Michael P. Snyder, Vadim N. Gladyshev, Tony Wyss-Coray, Kejun Ying

## Abstract

Human health is a single underlying state that no measurement observes directly. Diagnoses, blood tests, molecular profiles and images capture different facets at different times. Inferring health from such evidence requires a representation that integrates every modality, persists between observations and is revised as new evidence arrives. Here we introduce HealthFlux, a pan-modal world model that learns the latent dynamics of health from 4,920 features and three-dimensional brain and body MRI across eleven data domains in 502,166 UK Biobank participants. Its hybrid state-space architecture combines ODE-based evolution between observations with continuous-time recurrent updates when new measurements arrive. In held-out participants, recursive simulations without further observations predicted 1,256 diseases and death over twenty years, generally outperforming previously published state-of-the-art models. In four independent US cohorts, its performance was comparable to or better than that of these models. HealthFlux also exceeded specialized clinical risk scores for disease and mortality. HealthFlux predicts diseases excluded entirely from training, correctly ranking a future case above an unaffected person in 78% of comparisons, evidence that it has learned health itself rather than the diseases it was trained on. Medication-conditioned virtual clinical trials simulated drug-associated physiological changes and downstream disease risk. Simulated effects correlated with biomarker changes reported in 286 randomized trials and predicted disease and safety effects in 228 randomized trials withheld from model fitting. Each modality contributes information the others lack, and integrating them identifies individuals at risk whom single-modality models miss. HealthFlux treats health itself as the object of prediction through one continuously updated state, informed by any measurement, that supports disease-risk estimates years before diagnosis.

## Main Text

Clinical records and biological measurements capture different aspects of an individual’s health at different times. These observations reflect processes across molecular, physiological and organ systems.^1–3^ Epidemiological studies describe the co-occurrence and temporal ordering of diseases.^4–7^ The modelling problem is to infer a latent state that combines these partial observations, follows health between measurements and incorporates new evidence as it arrives. Population-scale biobanks now bring these observations together in the same participants.^8, 9^ Biological-age measures capture differences in health among individuals of the same chronological age.^10^ DNA-methylation biomarkers predict morbidity and mortality,^11–13^ while longitudinal molecular profiling and organ-specific analyses reveal distinct ageing trajectories across individuals and physiological systems.^14–16^ These measurements differ in timing and coverage. Blood tests may be repeated within months, MRI scans may be years apart, and diagnoses enter the record when disease is clinically recognized. An individual’s physiological state must be inferred from this incomplete and asynchronous evidence.

EHR models learn from sequences of clinical events,^17–21^ and multimodal approaches combine biological and clinical inputs for disease prediction.^22, 23^ A general latent-state representation must integrate information across these sources, retain it between observations and support predictions when disease-specific labels are scarce.

A world model represents a system through a latent state, simulates its dynamics and updates the state when new observations arrive.^24, 25^ This corresponds to the task of inferring an individual’s changing health from partial clinical, molecular, physiological and anatomical observations. Continuous-time latent-state methods describe change between irregular observations,^26–30^ and medical world models learn patient trajectories from longitudinal records.^31–33^ Combining simulation between observations with observation-driven revision allows a health representation to be queried between visits and updated as new information becomes available. Here we introduce HealthFlux, a pan-modal world model that learns the latent dynamics of health from clinical and biological observations in 502,166 UK Biobank participants. We test its predictive value in held-out participants and in four US cohorts—the Coronary Artery Risk Development in Young Adults (CARDIA) study, the Atherosclerosis Risk in Communities (ARIC) study, the Women’s Health Initiative (WHI) and the All of Us Research Program—and its generality through prediction of diseases excluded from training. Medication-conditioned state transitions are evaluated against observed biomarker responses and clinical-trial effects. Molecular and anatomical analyses examine the biological information used by the model across disease systems.

## Results

### Learning the latent dynamics of human health

HealthFlux integrated eleven data domains spanning longitudinal clinical records, demographics, genetics, blood biochemistry, haematology, physical function, lifestyle, proteomics, metabolomics, body MRI and brain MRI. The model combined 4,877 time-dependent clinical-event and numeric features, three-dimensional brain and body MRI volumes and 43 permanent-context variables (Fig. 1a). Each MRI volume was encoded by a three-dimensional convolutional encoder initialized from a pretrained MRI foundation model and fine-tuned jointly with the world model. The architecture followed the way health is observed in practice (Fig. 1b). Clinical evidence is sparse and irregular: a blood test may be repeated within months, an MRI scan may be acquired once, and a diagnosis is recorded only when disease is recognized, whereas the underlying physiology changes continuously in between. HealthFlux therefore carried each person’s health state forward in continuous time with a neural ordinary differential equation conditioned on age and permanent context, so that the state could be read at any age, including during long intervals without clinical contact. When a new diagnosis, measurement or image was recorded, a stochastic gate weighed it against the state expected from the person’s history, and a gated recurrent update revised the state, much as a clinician revises an assessment after a new finding. The deterministic and stochastic components, with 256 and 128 dimensions, formed one 384-dimensional health state (Fig. 1c), from which shared decoders estimated the risks of 1,256 diseases and death (Fig. 1d). An action branch, fitted with the world model held fixed, represented a medication by its pharmacological properties and shifted the health state after treatment started, so that futures with and without the medication could be simulated for the same individual (Fig. 1b,e). Ablations supported both temporal pathways (Figure S1). Removing the gated recurrent revision raised held-out next-event cross-entropy from 4.66 to 5.06 and lowered the mean five-year AUROC across the 1,256 diseases by 0.041, whereas removing ODE propagation left prediction after short intervals largely unchanged but increased the next-event timing error after five or more observation-free years from 875 to 1,203 days. Across internal widths from 64 to 1,024 units, held-out next-event cross-entropy and timing error were lowest at the width used in HealthFlux, and performance on the other metrics plateaued from widths of 256 to 384 onwards (Figure S2).

**Figure 1:**
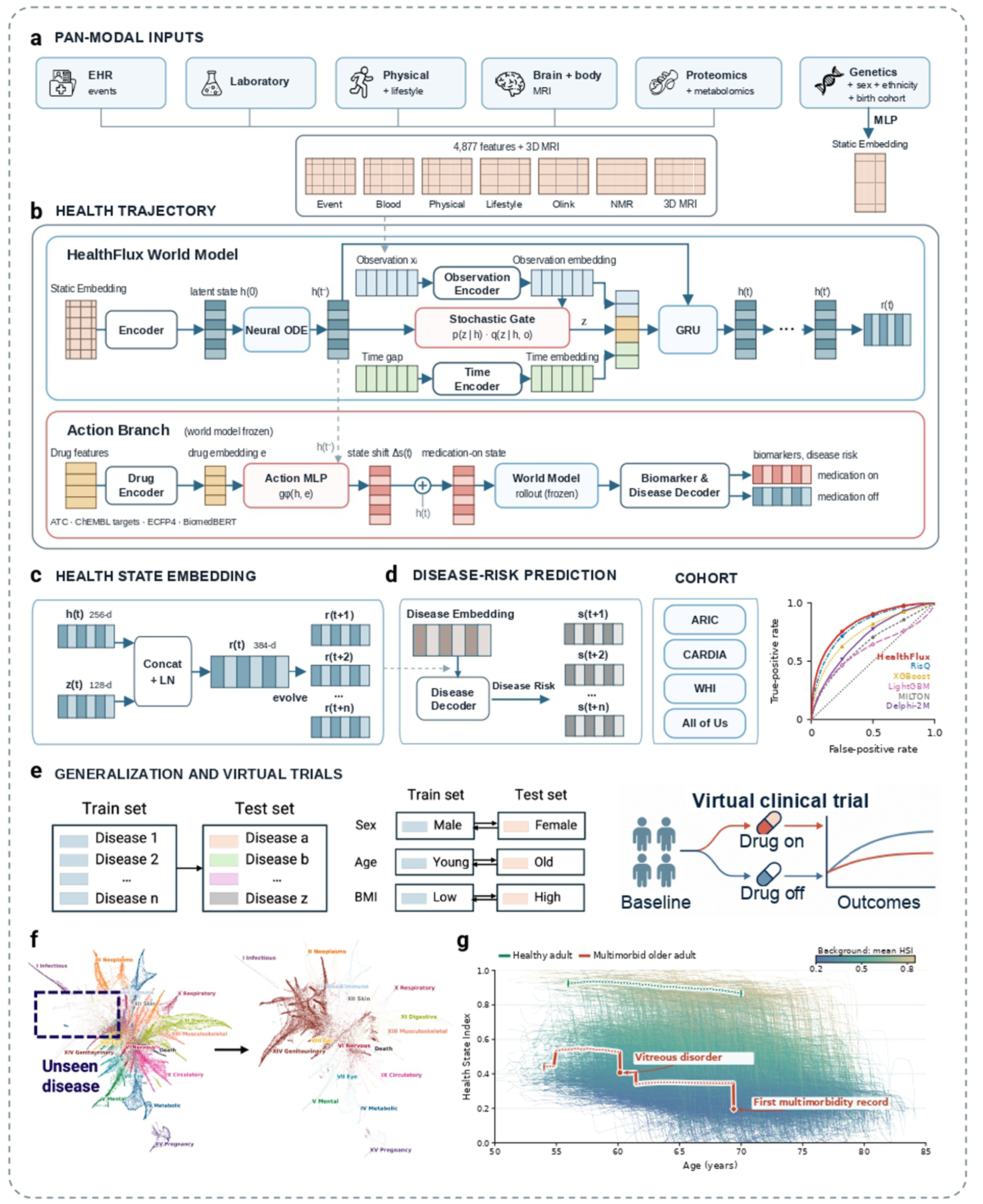
HealthFlux learns the latent dynamics of health from asynchronous pan-modal records. **a**, Pan-modal inputs across eleven data domains: 4,877 time-indexed clinical-event and numeric features, three-dimensional brain and body MRI volumes, and 43 permanent-context variables. **b**, HealthFlux world model: a neural ODE simulates the health state between observations, a stochastic gate and a GRU update revise it at each observation, and the action branch shifts it when a medication starts (world model frozen). **c**, Health state *r* (*t*), the layer-normalized concatenation of 256 deterministic and 128 stochastic dimensions. **d**, Multi-disease risk decoding, benchmark ROC curves in held-out UK Biobank participants and external evaluation in ARIC, CARDIA, WHI and All of Us. **e**, Generalization to unseen diseases and populations, and a schematic virtual clinical trial with paired medication-on and medication-off trajectories initialized from the same health state. **f**, Risk spectrum for a disease excluded from training. **g**, HSI trajectories of healthy and later-multimorbid individuals against the population background.

We used the latent state to predict future disease and mortality in held-out UK Biobank participants and evaluated disease prediction in four external cohorts (Fig. 1d). Disease prediction was also tested after target diagnoses were excluded from world-model and readout training (Fig. 1e,f). Separate experiments evaluated readouts on measurement identities or demographic groups withheld during readout fitting (Fig. 3c,e). These tests examined whether a state learned across conditions could support prediction with limited target-specific labels. Virtual clinical trial simulations compared paired medication-on and medication-off trajectories initialized from the same pre-medication state (Fig. 1e).

A separate Health State Index (HSI) summarized each state as a single number between 0 (death) and 1 (the health of long-term disease-free participants), using a readout trained on survival and disease burden. The same population-reference scale applied across participants and time points, allowing individual trajectories to be compared.

Individual trajectories illustrate how the model integrates information over time (Fig. 1g). One disease-free participant maintained a high HSI between ages 56 and 70. The participant who developed multimorbidity at age 69.4 already had a lower HSI by age 55, with a further decline after a vitreous-disorder record at about age 60.

Across held-out participants, the HSI at the baseline UK Biobank assessment declined with age and stratified later outcomes (Figure S3). Each 1-SD lower HSI was associated with a 1.67-fold higher hazard of death (95% CI 1.61–1.73) after adjustment for age and sex, and adding the HSI to age and sex increased the 10-year concordance for death from 0.708 to 0.726. The 10-year cumulative incidence of new multimorbidity fell from 71% in the lowest HSI decile to 11% in the highest; between these deciles, incidence differed 13-fold for type 2 diabetes and 17-fold for chronic obstructive pulmonary disease, but only 2-fold for cancer. Within each age band, the lowest HSI quintile had two to three times the 10-year mortality of the other quintiles. The HSI also agreed with participants’ own rating of their health, which the model did not receive: the median HSI was 0.45 among participants who rated their health as excellent and 0.24 among those who rated it as poor, an age-and sex-adjusted difference of 0.26 (95% CI 0.26–0.27). A lower HSI also preceded recorded chronic disease (Figure S4). Two years before the first record of 29 of the 31 chronic conditions that define multimorbidity, participants had a lower

HSI than matched controls of the same sex and age, most markedly before heart failure, chronic obstructive pulmonary disease and chronic kidney disease. For these conditions, the difference was already present ten years earlier and widened until the first record. Future cases of cancer, which this definition restricts to skin, breast and prostate cancer, had a slightly higher HSI than their controls.

### Predicting disease from a persistent and revisable latent state

HealthFlux captured age-associated patterns of recorded disease incidence across respiratory, reproductive, metabolic and cardiovascular conditions (Fig. 2a). Across 1,256 diseases and death, it achieved a five-year mean AUROC of 0.832 and average precision of 0.032 in held-out participants (Fig. 2b). The strongest evaluated comparator, RisQ,^23^ achieved a mean AUROC of 0.805 and average precision of 0.021. HealthFlux also exceeded XGBoost,^34^ LightGBM,^35^ MILTON^22^ and Delphi-2M^21^ on the same endpoints and risk sets (Table 1). Figure 2c shows disease-level five-year performance by disease system.

**Figure 2:**
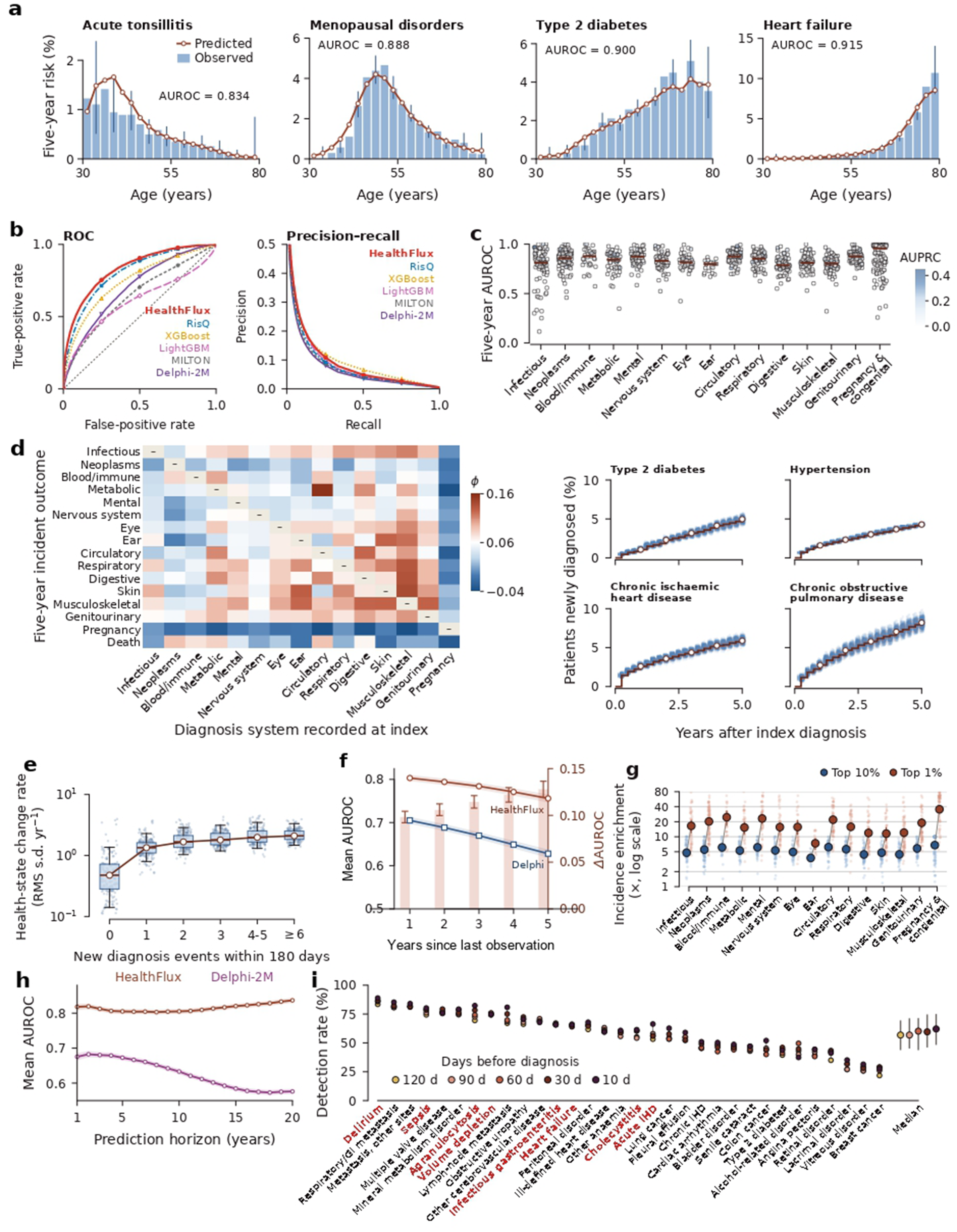
Disease prediction from a persistent and revisable health state. **a**, Observed incidence and predicted five-year risk by age for four diseases. **b**, Mean five-year ROC and precision–recall curves for 1,256 diseases and death. **c**, Disease-level AUROC by disease system (ICD-10 chapters; in **c** and **g**, Pregnancy & congenital combines the pregnancy, perinatal and congenital chapters); colour, AUPRC; bars, medians. **d**, Cross-system associations and four progression trajectories. **e**, State change rate by new diagnoses within 180 days. **f**, Mean AUROC after 1–5 years without new observations; bars, difference from Delphi-2M. **g**, Five-year incidence enrichment in top-risk groups. **h**, Cumulative/dynamic AUROC over 1–20 years; shading, 95% confidence intervals. **i**, Future cases with substantial prior morbidity flagged 10–120 days before diagnosis (32 diseases; acute conditions in red); right, median and interquartile range. IHD, ischaemic heart disease; GI, gastrointestinal.

**Figure 3:**
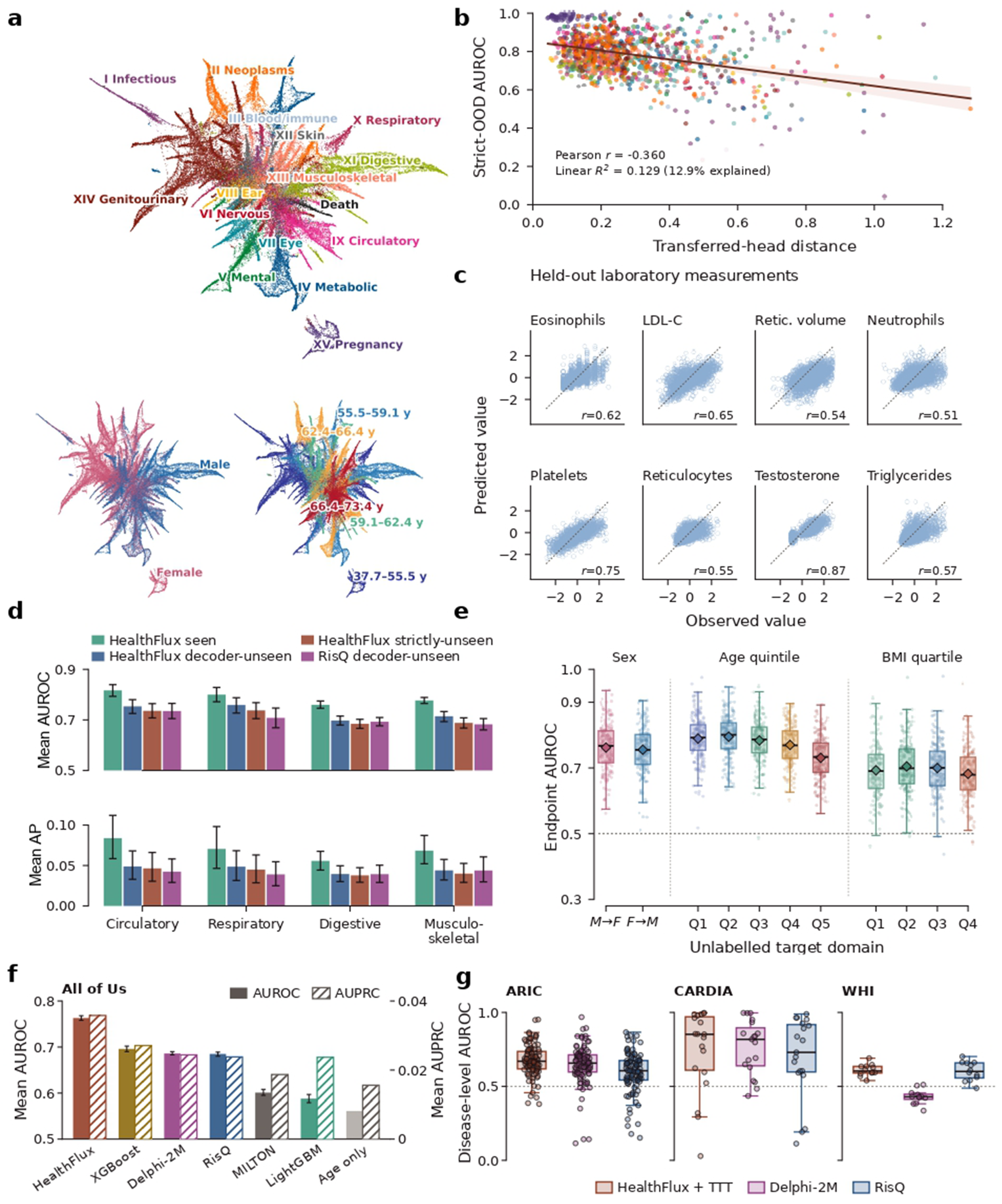
HealthFlux enables transfer across diseases, measurements and populations. **a**, Projection of risk profiles by dominant ICD-10 chapter, sex and age group. **b**, Strictly unseen-disease AUROC versus transferred-head distance across 1,256 diseases (Pearson *r* = −0.360; *R*^2^ = 0.129). **c**, Predicted versus observed repeat laboratory measurements withheld from readout training (pooled *r* = 0.629). **d**, Five-year mean AUROC and average precision for whole-chapter holdouts. **e**, Five-year readout transfer to a group withheld from readout fitting: M→F, fitted in men, tested in women; F→M, the reverse; landmark-age quintiles <47.9, 47.9–55.5, 55.5–61.3, 61.3–66.9 and ≥66.9 years; body-mass-index quartiles <24.6, 24.6–27.3, 27.3–30.5 and ≥30.5 kg/m^2^ (cut points from the training split). Diamonds and bars, mean AUROC with 95% bootstrap intervals. **f**, Zero-shot mean AUROC (solid) and AUPRC (hatched) in All of Us, 971 common outcomes. **g**, External-cohort generalization: disease-level AUROC in ARIC, CARDIA and WHI for HealthFlux with label-free test-time training (HealthFlux+TTT), Delphi-2M and RisQ. In **e** and **g**: points, diseases; boxes, medians and interquartile ranges; whiskers (**g**), 1.5 times that range.

**Table 1:** Disease prediction and transfer evaluations.

**a, Specific disease prediction**
| Model | Native score or representation | AUROC↑ | AUPRC↑ |
| --- | --- | --- | --- |
| *Composite cardiovascular disease (n = 40,712; 267 events)* |  |  |  |
| QRISK3 | 10-year CVD risk | 0.743 | 0.019 |
| SCORE2 | 10-year fatal/non-fatal CVD risk | 0.728 | 0.017 |
| PREVENT | 10-year ASCVD risk | 0.733 | 0.018 |
| Framingham | 10-year general CVD risk | 0.707 | 0.017 |
| **HealthFlux** | **Shared latent state** | **0.776** | **0.026** |
| *All-cause death (n = 63,000; 229 events)* |  |  |  |
| Charlson | Age-adjusted comorbidity index | 0.754 | 0.018 |
| Elixhauser | van Walraven mortality score | 0.667 | 0.015 |
| **HealthFlux** | **Shared latent state** | **0.846** | **0.081** |

**b, General disease prediction and OOD generalization**
| Task | Metric | Machine learning |  |  | Deep/foundation |  | World model |
| --- | --- | --- | --- | --- | --- | --- | --- |
|  |  | XGBoost | LightGBM | MILTON | Delphi-2M | RisQ | **HealthFlux** |
| General disease prediction | AUROC↑ | 0.746 | 0.604 | 0.647 | 0.699 | 0.805 | **0.832** |
| (1,256 diseases and death) | AUPRC↑ | 0.019 | 0.016 | 0.010 | 0.012 | 0.021 | **0.032** |
| Strictly unseen disease | AUROC↑ | N/A | N/A | N/A | N/A | N/A | **0.783** |
|  | AUPRC↑ | N/A | N/A | N/A | N/A | N/A | **0.017** |
| Unseen population (sex) | AUROC↑ | 0.761 | 0.732 | 0.627 | N/A | N/A | **0.790** |
| (2 directional settings) | AUPRC↑ | 0.058 | 0.055 | 0.033 | N/A | N/A | **0.072** |
| Unseen population (age) | AUROC↑ | 0.752 | 0.733 | 0.587 | N/A | N/A | **0.787** |
| (5 quintile settings) | AUPRC↑ | 0.053 | 0.049 | 0.029 | N/A | N/A | **0.069** |
| Unseen population (BMI) | AUROC↑ | 0.706 | 0.680 | 0.584 | N/A | N/A | **0.708** |
| (4 quartile settings) | AUPRC↑ | 0.071 | 0.069 | 0.050 | N/A | N/A | **0.071** |
| Unseen measurement | Pearson $r$ ↑ | N/A | N/A | N/A | N/A | N/A | **0.629** |
| (standardized values) | MSE↓ | N/A | N/A | N/A | N/A | N/A | **0.574** |

Observed disease sequences included type 2 diabetes followed by chronic kidney disease and chronic ischaemic heart disease followed by heart failure (Fig. 2d). These sequences are consistent with clinical descriptions of multimorbidity and disease progression.^4–6^

The inferred state responded to new clinical information. Participants with six or more new diagnoses over 180 days had a 4.4-fold greater rate of state change than participants with no new diagnoses (Fig. 2e). Endpoint-level agreement between predicted risk and observed incidence across EHR-history strata provided a complementary cross-sectional check (Figure S5). We simulated the latent state for one to five years while withholding all subsequent EHR events and biological observations. At each query, the model predicted outcomes during the following five years, so the last outcome window covered years 5–10 after the last observation. The mean-AUROC advantage over Delphi-2M increased from 0.098 at a one-year gap to 0.128 at a five-year gap (Fig. 2f). Participants with the highest predicted risks had greater observed disease incidence. Across disease systems, mean five-year incidence enrichment ranged from 3.8-fold to 6.8-fold in endpoint-specific top-10% groups and from 7.4-fold to 35.5-fold in top-1% groups (Fig. 2g). For the median disease, the 10% of participants with the highest predicted risk accounted for 51% of all new cases over five years, and their five-year risk was 9.5 times that of the remaining 90%. For heart failure, 29% of the top 1% were diagnosed within five years, compared with 1.4% of all participants at risk, and the top 10% contained 72% of new cases; for type 2 diabetes, the corresponding figures were 51% (300 of 585) versus 2.6% and 68%, and for chronic ischaemic heart disease 51% versus 3.3% and 55%. Of participants in the top 1% of predicted mortality risk, 74% died within five years, compared with 4.2% overall.

For longer-range prediction, HealthFlux recursively sampled the next diagnosis and its timing, evolved the health state, and fed each generated event back before continuing, without access to observed post-landmark events. Its equal-endpoint mean AUROC remained between 0.803 and 0.837 across annual horizons from one to twenty years and exceeded Delphi-2M throughout, including 0.837 versus 0.576 at twenty years (Fig. 2h). At twenty years, HealthFlux thus ranked a person who developed a given disease above one who did not in 84 of 100 random pairs, compared with 58 of 100 for Delphi-2M. In a separate retrospective analysis of first recorded diagnoses, 32 diseases were selected from 1,256 using validation data only (Figure S6a,b). The median disease-level detection rate among high-history-burden future cases ranged from 56.8% at 120 days to 62.1% at 10 days before diagnosis (Fig. 2i). The corresponding full-case medians were 29.9% and 38.3%, and detection was higher in the high-history-burden group in 150 of 160 disease–lead-time pairs (Figure S6c,d). The alert thresholds were set in validation data so that 5% of people without the disease would be flagged at a routine visit; at these thresholds, HealthFlux flagged a median of 57% of future cases with extensive prior morbidity four months before diagnosis. Early warning was strongest for conditions that typically present acutely, often as emergency admissions (highlighted in Fig. 2i): ten days before diagnosis, a median of 71% of their future cases with extensive prior morbidity were flagged, compared with 56% for the other diseases. HealthFlux flagged 89% of these patients who later developed delirium, 82% for agranulocytosis, 80% for sepsis, 75% for volume depletion, 66% for acute ischaemic heart disease and 66% for heart failure; across all patients, the corresponding figures were 70%, 70%, 55%, 51%, 41% and 48%. For acute events such as sepsis or decompensated heart failure, an alert ten days or more before hospital presentation leaves time for earlier assessment and treatment.

### Testing the transferability of the learned latent state

Individual disease-risk profiles were organized by disease system, sex and age in the two-dimensional projection (Fig. 3a). We tested whether the learned state supported predictions when target-specific information was withheld from training. All analyses used HealthFlux states and setting-specific semantic readouts; strict disease transfer additionally retrained the world model after excluding the held-out diagnoses.

For disease transfer, a semantic readout combined the frozen latent state with a text embedding of the queried disease description. Diagnosis-exclusion experiments tested whether this readout could predict targets withheld from training. In the decoder-unseen setting, target disease labels were withheld from disease-readout fitting. In the strict setting, target diagnoses were also excluded from world-model inputs, training targets and output support, and the model was retrained under these exclusions. Strict transfer achieved a mean AUROC of 0.783 across 1,256 diseases (Table 1). Whole-chapter exclusions extended this test across circulatory, respiratory, digestive and musculoskeletal diseases. Across all four chapters, AUROC decreased as target diagnoses were excluded first from readout fitting and then from world-model training. HealthFlux achieved higher point estimates than RisQ in the matched decoder-unseen comparison (Fig. 3d). The cosine distance between transferred and supervised risk directions for the same disease was associated with strict-transfer AUROC (Pearson *r* = −0.360; *R*^2^ = 0.129; Fig. 3b). In practical terms, for a disease that the model had never seen during training, a person who went on to develop it was assigned a higher risk than a person who did not in 78 of 100 randomly drawn pairs, compared with 83 of 100 for diseases with training labels. This corresponds to clinical situations in which labelled outcomes are not available for model training, such as a rare disease with too few recorded cases or a newly defined condition; HealthFlux provided risk estimates for such diseases from their written description and the learned health state alone.

We tested a second form of transfer by withholding assay identities from repeat-measurement readout fitting. The readout combined the source-visit state, current measurement and elapsed time to predict the next recorded value. For the eight laboratory identities shown in Fig. 3c, pooled correlation was 0.629 and mean squared error was 0.574, compared with 0.679 for carrying the current value forward. We also fitted disease readouts with labels from selected demographic groups and evaluated them in held-out sex, age or BMI groups. AUROCs across these settings were approximately 0.6–0.8 (Fig. 3e). Transfer AUROC was lower in the oldest age quintile and highest BMI quartile (Fig. 3e).

External-cohort transfer was tested in CARDIA, ARIC and WHI using label-free test-time training (HealthFlux+TTT). Adaptation updated normalization parameters from pre-landmark histories without outcome labels, while retaining the trained disease decoder. Median disease-level AUROCs were 0.852 in CARDIA, 0.670 in ARIC and 0.604 in WHI across each cohort’s common evaluable endpoints. HealthFlux+TTT achieved higher median AUROC than the evaluated Delphi-2M and RisQ pipelines in each cohort (Fig. 3g). We further tested zero-shot prediction in All of Us, a US cohort in which diagnosis history, age, sex, clinical laboratory tests, physical measurements and proteomics were available. Across the 971 outcomes evaluable for all models, HealthFlux had the highest mean AUROC (0.763, 95% CI 0.758–0.768), compared with 0.696 for XGBoost, 0.686 for Delphi-2M, 0.685 for RisQ, 0.601 for MILTON and 0.588 for LightGBM, and 0.561 for age alone. It also had the highest mean AUPRC (0.036, versus 0.019–0.027 for the other models and 0.016 for age alone). HealthFlux exceeded each comparator by 0.067–0.175 in mean outcome-level AUROC and had the higher AUROC for 93.6–97.4% of outcomes (Fig. 3f).

Table 1 summarizes these prediction and transfer results. In the endpoint-matched comparisons, HealthFlux also achieved higher AUROC and average precision than the clinical risk scores evaluated for cardiovascular disease and all-cause death (Table 1a).

### Simulating the effects of medications on physiology and disease risk

We examined whether HealthFlux could simulate the effects of medications on physiology and disease risk. Starting a medication was represented as an action that alters how the latent health state evolves, and biomarker and disease-risk trajectories were compared between paired simulations with and without the medication, both starting from the same pre-medication state. We first considered statins and antihypertensive drugs, two classes with well-characterized clinical effects. Following statin initiation, the medication-on trajectories showed lower LDL cholesterol and apolipoprotein B, a more favourable health state and lower risks of chronic ischaemic heart disease and stroke than the corresponding medication-off trajectories. Antihypertensive initiation similarly lowered systolic and diastolic blood pressure, maintained a more favourable health state and attenuated the accumulation of stroke and heart-failure risk. Observed biomarker measurements were consistent with the medication-on trajectories (Fig. 4a).

**Figure 4:**
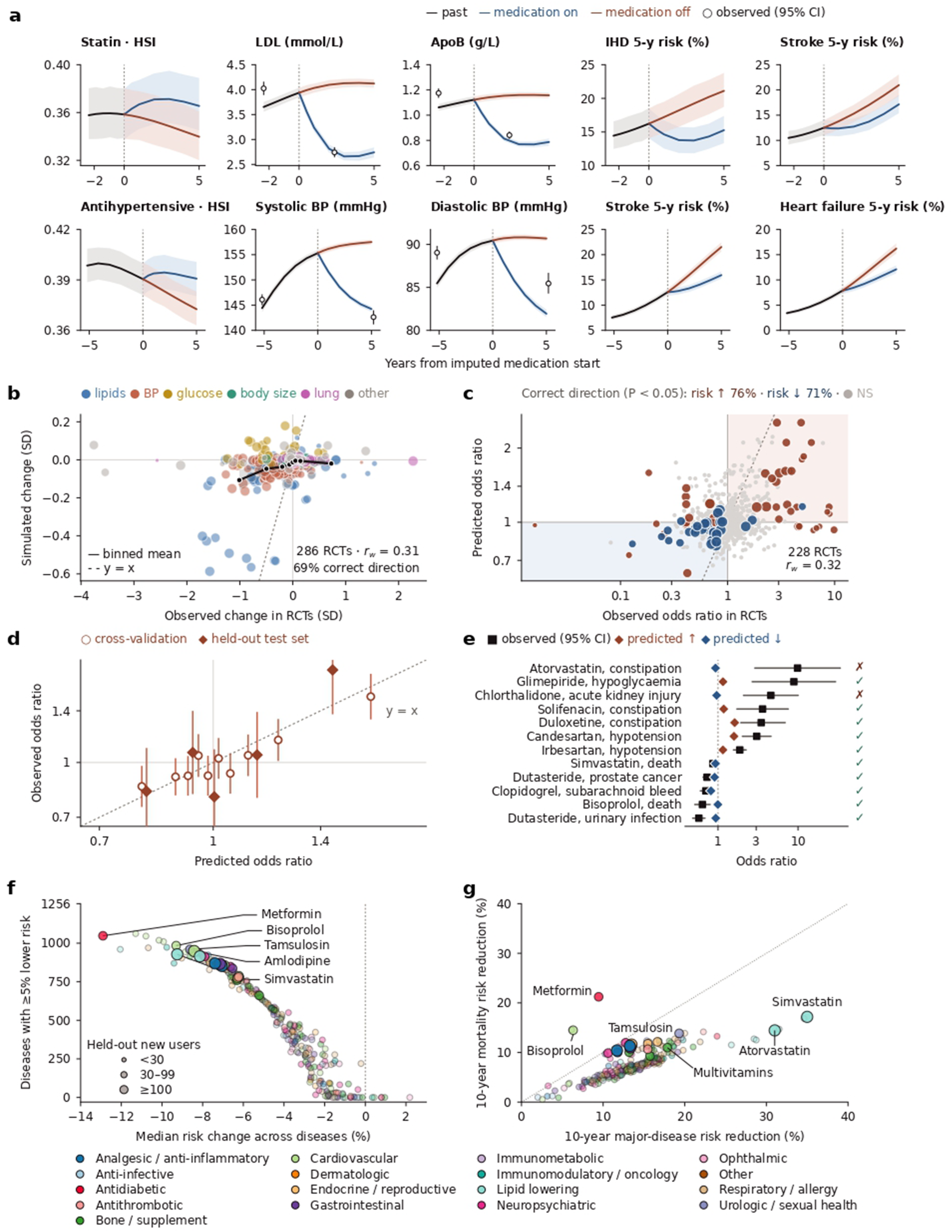
HealthFlux simulates medication effects on physiology and disease risk. **a**, Simulated trajectories with (blue) and without (rust) statin or antihypertensive treatment from the same pre-treatment state; circles show observed measurements. **b**, Simulated versus reported biomarker changes in 286 randomized controlled trials (RCTs). **c**, Predicted versus observed odds ratios for 655 outcomes in 228 RCTs withheld from model fitting. **d**, Calibration of predicted odds ratios. **e**, Outcomes with the strongest trial evidence. **f**, Predicted disease-risk reductions for 259 medications across 1,256 diseases. **g**, Predicted 10-year reductions in major-disease and mortality risk in people free of major disease. *r_w_*, precision-weighted correlation. Simulated effects are model predictions and do not represent individual causal treatment effects.

To test whether medication effects learned from routine records agree with randomized evidence across many drugs, we represented each drug by its therapeutic class, molecular targets, chemical structure and mechanism of action, which allowed information to be shared across related drugs.

We then emulated randomized controlled trials (RCTs) that reported biomarker changes relative to placebo or a shared background therapy. Both arms of each trial were simulated from the same baseline states of trial-eligible, held-out UK Biobank participants, and the simulated between-arm difference was compared with the reported difference. Across 350 drug–biomarker pairs from 286 RCTs, covering 96 drugs and 14 biomarkers, simulated and reported changes were correlated (precision-weighted *r* = 0.31, 95% CI 0.11–0.49), and the direction of change agreed for 69% of the 124 pairs with a significant change in the trials (*P* < 0.05; Fig. 4b). Simulated changes were smaller than reported changes, so this agreement reflected their relative ordering rather than their absolute magnitude. Within UK Biobank, simulated changes also agreed with the changes observed in held-out new users of 81 drugs (precision-weighted *r* = 0.64); agreement with trials was highest for lipids and blood pressure, and the results were robust to how trial data were extracted (Figure S7).

Effects on disease are harder to learn from routine records, because people who start a medication often differ in health from those who do not (confounding by indication). We therefore predicted the effect of a drug on each outcome by combining its HealthFlux drug and disease representations with the trial design and the effects reported in other trials, and evaluated these predictions on RCTs withheld entirely from model fitting. Across 655 drug–outcome comparisons from 228 RCTs comparing a drug with placebo or with no added drug, covering 98 drugs and 79 outcomes that included target diseases, adverse events and death, predicted and observed odds ratios were correlated (precision-weighted *r* = 0.32, 95% CI 0.23–0.41). Among the 84 comparisons with a significant effect in the trial (*P* < 0.05), the direction was predicted correctly for 76% of increases in risk and 71% of decreases (Fig. 4c). Observed effects increased with predicted effects and lay close to the line of identity, both in cross-validation and in a sealed test set of 46 RCTs that was evaluated once (Fig. 4d). In this test set, the correlation was 0.46 (95% CI 0.24–0.63), compared with 0.12 (−0.06 to 0.34) for extrapolation from pooled estimates of the same drug and outcome in other trials (difference 0.34, 95% CI 0.11–0.52). Of the twelve outcomes with the strongest trial evidence, ten were predicted in the correct direction, including more hypoglycaemia with glimepiride, more hypotension with candesartan and irbesartan, and fewer deaths with simvastatin and bisoprolol (Fig. 4e).

Together, these results indicate that the physiological effects of medications can be learned from routine longitudinal records, whereas their effects on disease require anchoring in randomized evidence. We combined both principles to simulate the long-term consequences of individual medications. For each medication, the simulated action was learned from the biomarker changes observed in its new users, and for three well-established pathways—low-density lipoprotein (LDL) cholesterol, systolic blood pressure and glycaemia—the link between biomarker change and disease risk was calibrated to effects per unit change reported in trial meta-analyses.

We then screened 259 UK Biobank medication codes across 1,256 diseases, simulating each medication in held-out participants who started it. The predicted breadth of effect differed markedly among medications (Fig. 4f). Among medications with at least 30 held-out new users, metformin lowered the predicted risk of 1,048 diseases by at least 5%, and bisoprolol, tamsulosin, amlodipine and simvastatin each lowered the predicted risk of more than 900 diseases (Table 2). The broadest predicted benefits were concentrated among cardiovascular and metabolic therapies but also included tamsulosin, suggesting that some medications may influence multiple disease risks through broader changes in health state. For these candidates, predicted reductions spanned all 14 disease systems and were largest for circulatory disease (Figure S8a). These estimates are predictions rather than evidence that the medications prevent each individual disease. In a shared reference cohort free of major disease, medications also showed distinct predicted 10-year profiles. Statins produced the largest reductions in major-disease risk; simvastatin was associated with predicted reductions of approximately 35% in major-disease risk and 17% in mortality risk. Metformin produced a smaller reduction in major-disease risk but the largest reduction in mortality risk, approximately 21%, and bisoprolol showed the same pattern of a larger reduction in mortality than in major-disease risk (Fig. 4g; Table 2). Screen estimates were reproducible across independently trained models (Pearson *r* ≥ 0.91 between random seeds; Figure S8b,c). These estimates identify candidates for testing in drug-repurposing and healthy-ageing studies.

**Table 2:** Drug candidates from medication-conditioned simulations.

|  | Simulated 10-year risk reduction |  |  |
| --- | --- | --- | --- |
| Medication / UKB code | Mortality (%) | Major disease (%) | Diseases with $\geq 5\%$ lower risk / 1,256 |
| A. Mortality-risk reduction exceeds major-disease-risk reduction |  |  |  |
| Metformin<br>1140884600 | 21.2 | 9.5 | 1,048 |
| Bisoprolol<br>1140879760 | 14.5 | 6.3 | 984 |
| B. Major-disease-risk reduction exceeds mortality-risk reduction |  |  |  |
| Simvastatin<br>1140861958 | 17.2 | 35.0 | 928 |
| Atorvastatin<br>1141146234 | 14.4 | 31.0 | 915 |
| Tamsulosin<br>1140926934 | 13.9 | 19.3 | 960 |
| Multivitamins<br>1140852976 | 11.0 | 17.9 | 760 |
| C. Broad disease-risk signals across more than 900 diseases |  |  |  |
| Metformin<br>1140884600 | 21.2 | 9.5 | 1,048 |
| Bisoprolol<br>1140879760 | 14.5 | 6.3 | 984 |
| Tamsulosin<br>1140926934 | 13.9 | 19.3 | 960 |
| Amlodipine<br>1140879802 | 12.4 | −1.3 | 949 |
| Simvastatin<br>1140861958 | 17.2 | 35.0 | 928 |

### Integrating modality-specific disease signals

The modality analyses in Fig. 5 used HealthFlux states and prespecified disease readouts. The world model’s learned event embeddings organized clinically related endpoints in disease space (Fig. 5a). Modality-attribution maps showed that metabolomic, proteomic and imaging inputs emphasized different regions of this space. Relative attribution was greater for blood bio-chemistry, metabolomics and body MRI in metabolic-disease scores, proteomics in blood-and immune-disorder scores, lifestyle in mental-disorder scores, and physical function in circulatory-disease scores (Fig. 5b). Across 3,567 measured features, similar disease-response profiles connected molecular and physiological measurements within and across modalities (Fig. 5c). Removing each input modality changed the model’s predictions, with the largest root-mean-square score change for proteomics (Fig. 5d). Protein-derived and metabolite-derived disease similarities were strongly correlated (Pearson *r* = 0.856; Fig. 5e).

**Figure 5:**
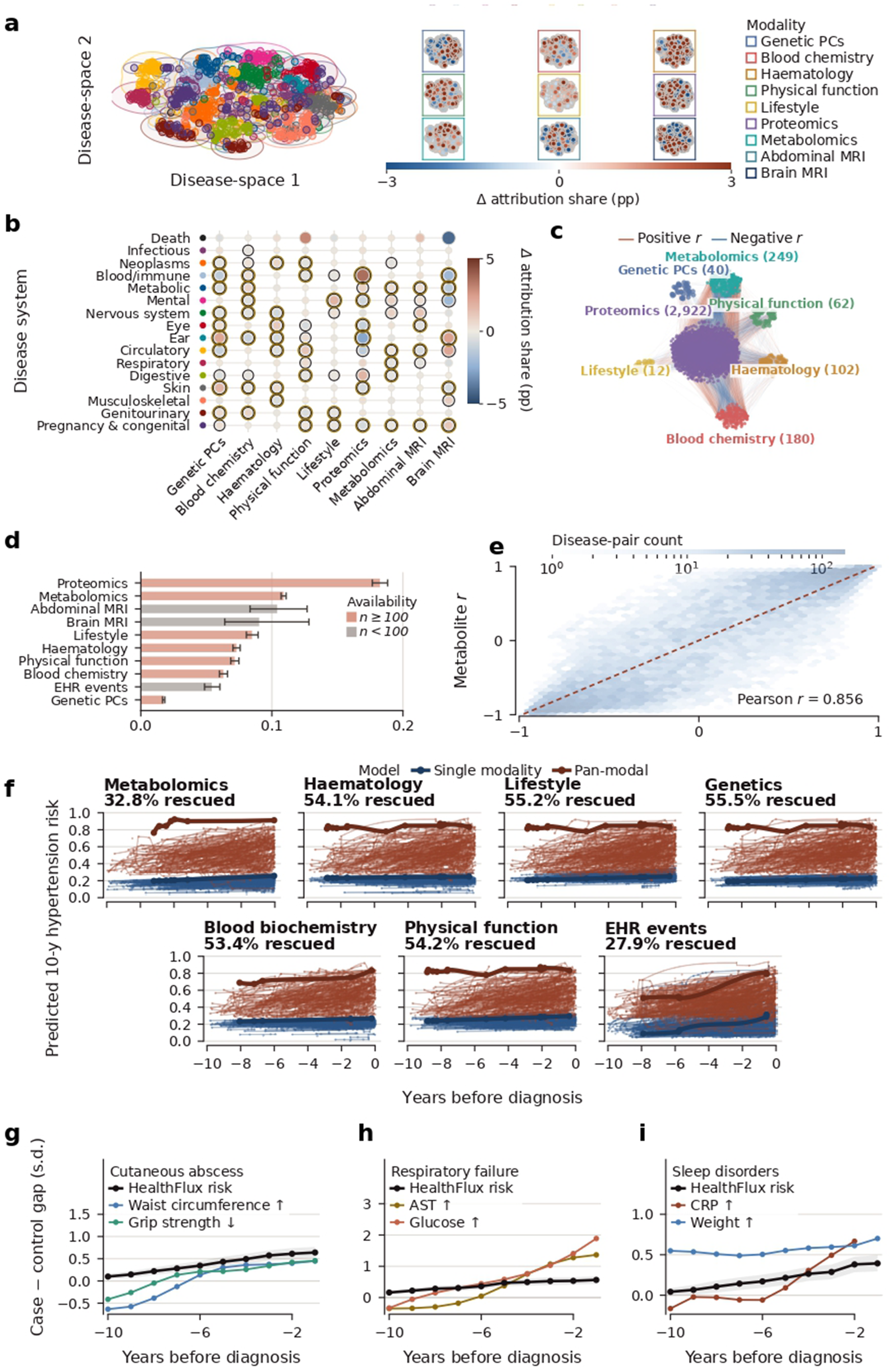
Pan-modal organization of the shared health state. **a**, Disease space coloured by disease system, with modality attribution maps; **b**, modality attribution by disease system; **c**, cross-modal feature network; **d**, modality ablation (EHR, electronic health record); **e**, protein–metabolite concordance; **f**, pan-modal rescue of hypertension cases; **g–i**, prediagnostic trajectories (AST, aspartate aminotransferase; CRP, C-reactive protein).

Combining modalities also identified future hypertension cases missed by single-modality models (Fig. 5f). We selected participants whose first hypertension record occurred between ages 65 and 75 and whose age-65 score was below the relevant single-modality threshold at nominal 90% specificity. Using the pan-modal threshold fixed in validation data at nominal 90% specificity, HealthFlux identified 27.9–55.5% of the cases missed by the corresponding single-modality model.

HealthFlux risk trajectories in Fig. 5g–i were used for matched case–control comparisons of predicted risk and biological measurements at annual landmarks before diagnosis. As diagnosis approached, risk gaps increased alongside shifts toward greater waist circumference and lower grip strength in future cutaneous-abscess cases, higher glucose and aspartate aminotransferase in future respiratory-failure cases, and higher C-reactive protein and body weight in future sleep-disorder cases relative to matched controls. These comparisons related predicted risk to biological differences that developed before disease was recorded.

### Relating structural imaging to disease risk

Structural brain and body MRI provide an anatomically grounded test of the shared latent state learned by HealthFlux within the UK Biobank imaging resource.^9, 36, 37^ We examined whether this state retained stable, disease-specific brain patterns and captured associations between brain structure and disease systems. Across ten MRI-derived measures, advancing age was associated with lower grey-matter volume, cortical thickness and selected regional volumes, alongside higher cerebrospinal-fluid fraction and ventricular volume.^38, 39^ Over the same age range, the rate of newly recorded diagnoses also increased (Fig. 6a). Previous studies have linked brain age to mortality and vascular risk factors to structural brain differences.^40, 41^

**Figure 6:**
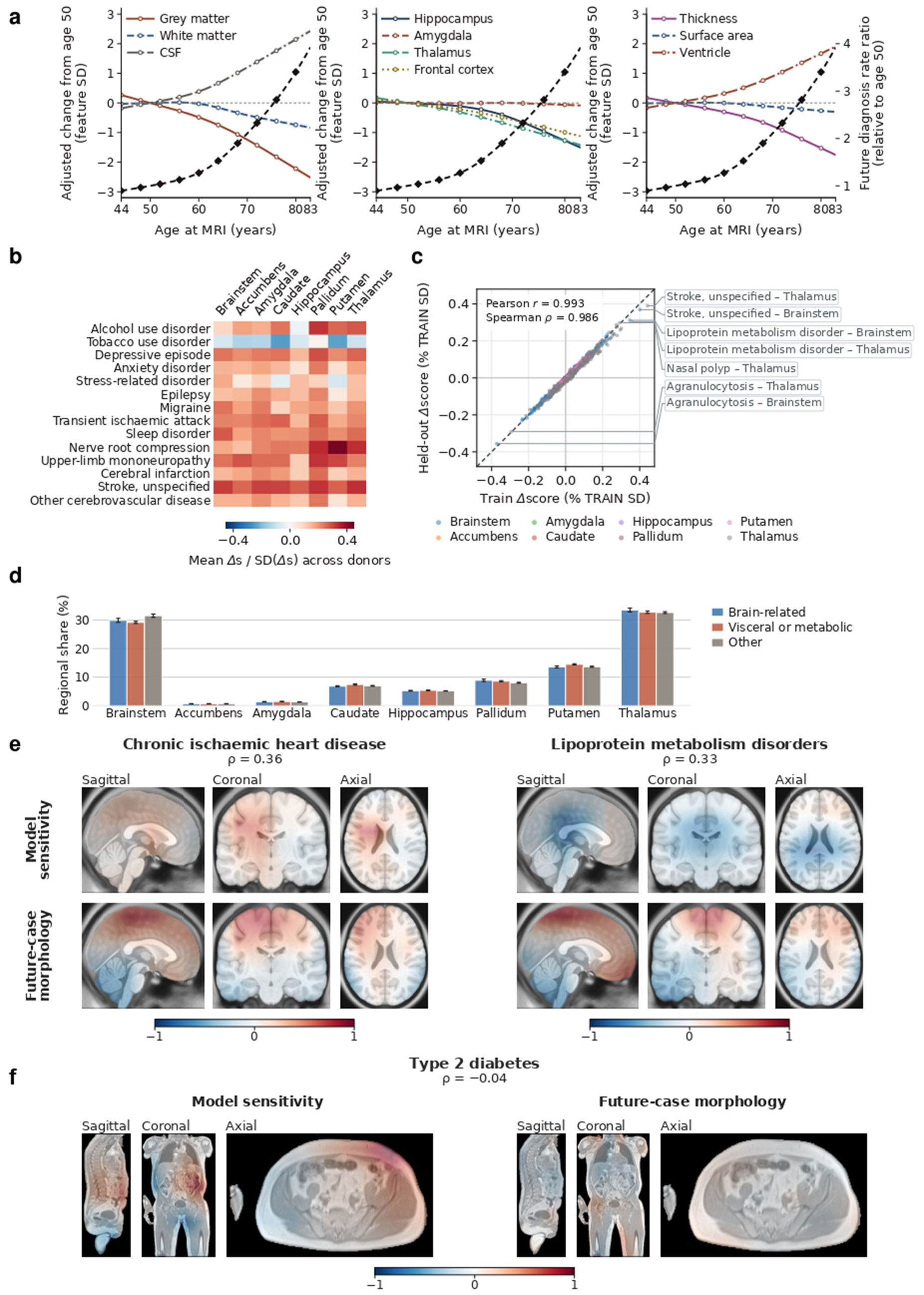
HealthFlux relates structural imaging to disease risk. **a**, Age trajectories of ten brain-MRI measures and the future-diagnosis rate ratio. **b**, Regional-ablation sensitivity for 14 brain-related diseases. **c**, Training-versus-held-out disease–region sensitivities (Pearson *r* = 0.993; *n* = 1, 560; labels, disease–region pairs). **d**, Regional contribution shares by disease group. **e**, **f**, Brain-MRI (**e**) and body-MRI (**f**) sensitivity and future-case morphology for chronic ischaemic heart disease and lipoprotein metabolism disorders (spatial Spearman *ρ* = 0.36, 0.33) and type 2 diabetes (*ρ* = −0.04); maps scaled to the 99th-percentile absolute value and centred at zero.

Replacing eight FIRST-segmented subcortical structures in the brain MRI with their population-mean intensity produced disease-specific changes in predicted risk across 14 brain-related diseases (Fig. 6b). For these diseases, replacement predominantly lowered predicted risk (102 of 112 disease–region pairs, 91%, versus 59% across all 195 diseases; nicotine dependence was the main exception), indicating that participant-specific subcortical appearance raised the predicted risk of neurological, psychiatric and cerebrovascular disease. Across 1,560 disease–region pairs, regional prediction sensitivities were consistent between training and held-out UK Biobank participants (Pearson *r* = 0.993; Fig. 6c). The thalamus, brainstem and putamen, the three largest structures, accounted for most of this sensitivity, with similar regional profiles for neurological, psychiatric, visceral, metabolic and other systemic diseases (Fig. 6d).

We next asked whether the image regions that drove disease predictions coincided with anatomical differences already present before diagnosis. For brain MRI, we examined chronic ischaemic heart disease and lipoprotein metabolism disorders, two cardiometabolic conditions whose risk factors are associated with brain structure;^41^ for body MRI, we examined type 2 diabetes, for which abdominal adiposity is an established risk factor. In both brain examples, model sensitivity was positively correlated with the prediagnostic differences between future cases and controls, most clearly for chronic ischaemic heart disease (Spearman *ρ* = 0.36; 95% bootstrap interval 0.20–0.39) and with a similar coefficient for lipoprotein metabolism disorders (*ρ* = 0.33; −0.09 to 0.42; Fig. 6e). HealthFlux therefore drew on brain regions that already differed before these systemic diseases were diagnosed. By contrast, body-MRI sensitivity for type 2 diabetes showed little correspondence with prediagnostic differences (*ρ* = −0.04; −0.18 to 0.10; Fig. 6f); together with the contributions of blood biochemistry, metabolomics and body MRI to metabolic-disease scores (Fig. 5b), this suggests that type 2 diabetes was predicted from information distributed across modalities rather than from body anatomy alone. Each comparison included three to five prediagnostic validation cases.

## Discussion

HealthFlux models human health as an evolving latent state inferred from clinical events, laboratory and physical measurements, lifestyle, molecular profiles and imaging. It learns dynamics that allow the state to be simulated between observations and revised as new evidence arrives, and a single state supports risk estimates for 1,256 diseases and death.

In clinical terms, the main results can be stated as risks and proportions. For the median disease, the 10% of held-out participants whom HealthFlux ranked at highest risk accounted for 51% of new cases over the following five years and were 9.5 times as likely to develop the disease as the remaining 90%. Among the 1% at highest predicted risk, 29% were diagnosed with heart failure within five years, compared with 1.4% of all participants, and 74% died, compared with 4.2%. The model retained this ability without new information: simulated forward for twenty years without further observations, it ranked a person who developed a disease above one who did not in 84 of 100 random pairs, compared with 58 of 100 for Delphi-2M. At alert thresholds that would flag 5% of people without the disease at a routine visit, it flagged a median of 57% of future cases with extensive prior morbidity four months before diagnosis. In randomized trials withheld from model fitting, it predicted the direction of 76% of significant increases and 71% of significant decreases in disease and safety outcomes, and integrating modalities identified 28–56% of future hypertension cases missed by single-modality models.

Each diagnosis can also be placed within a longer model-inferred trajectory. The Health State Index (HSI), a single score between 0 (death) and 1 (the health of long-term disease-free participants) read from the health state, summarizes these trajectories on a common scale. Each 1-SD lower HSI at baseline was associated with a 1.67-fold higher hazard of death after adjustment for age and sex, and the 10-year incidence of new multimorbidity was 71% in the lowest HSI decile and 11% in the highest. Two years before the first record of 29 of 31 chronic conditions, future cases had a lower HSI than matched controls of the same sex and age (Figure S4). For heart failure, the future case had the lower HSI in 70% of case–control pairs, and the mean HSI of future cases was already 17% lower than that of controls ten years before diagnosis (0.31 versus 0.37). A diagnosis can therefore be placed within a process already under way, and following these trajectories allows gradual changes between observations to be examined alongside changes accompanying newly recorded illness.

Learning across diseases is particularly useful when target-specific labels are scarce: for rare diseases with too few recorded cases to train a dedicated model, for conditions that are newly defined, such as post-COVID-19 condition, which had no dedicated diagnosis code before the pandemic, and for health systems in which an outcome is seldom coded. HealthFlux predicted disease risk after target diagnoses were removed from both world-model and readout training, ranking a future case above a non-case in 78 of 100 pairs, close to the 83 of 100 achieved with full training labels. This transfer indicates that the learned state contains predictive information shared across diagnostic categories. The assay and demographic experiments reused the same representation, with assay identities and group-specific disease labels withheld at the readout-fitting stage, and the resulting readouts predicted repeat measurements and disease risk in these held-out settings.

The molecular and imaging analyses gave biological content to this evolving state. Proteomic, metabolomic, physiological and anatomical measurements contributed distinct disease signals, and similar disease-response profiles connected measurements within and across modalities. Subcortical structures most consistently raised the predicted risk of brain-related diseases. Brain-MRI sensitivity aligned with anatomical differences preceding cardiometabolic diagnoses, whereas type 2 diabetes prediction drew on information beyond body anatomy.

An evolving latent state brings measurements collected at different times into a common representation for disease prediction. Our evaluations used diagnoses as recorded in health records, which lag disease onset and depend on contact with health care, and prediction in CARDIA, ARIC and WHI used adaptation to unlabelled pre-landmark histories. Prospective studies and independent cohorts with repeated molecular measurements and imaging can test how these dynamics relate to functional decline and the onset of disease, and whether updating the state improves clinical decisions as new observations arrive.

## Data Availability

Participant-level data used in this study are held by UK Biobank, CARDIA, ARIC, WHI and the All of Us Research Program. Access is governed by each cohort's application and data-use requirements. UK Biobank data are available to approved researchers through its data-access process (https://www.ukbiobank.ac.uk/use-our-data/). CARDIA and ARIC data can be requested through NHLBI BioLINCC (https://biolincc.nhlbi.nih.gov/). WHI data access is described at https://www.whi.org/md/access-to-whi-data. All of Us data are available to approved researchers through the Researcher Workbench (https://www.researchallofus.org/).

## Methods

### Study population, governance and data split

We used de-identified data from 502,166 UK Biobank participants with linked health records and biological phenotyping.^8^ UK Biobank operates under research-tissue-bank approval from the North West Multi-centre Research Ethics Committee (11/NW/0382 and subsequent renewals), and all participants provided written informed consent. We analysed hospital diagnoses, mortality, blood measurements, physical and lifestyle traits, plasma proteomics, NMR metabolomics, brain and body MRI, genetic principal components and demographic variables. We represented diagnoses at the three-character ICD-10 level. The disease panel comprised 1,256 diseases and death. We removed records after recorded death and treated death as an absorbing event.

We assigned participants before model development to non-overlapping training (*n* = 351, 219), validation (*n* = 75, 629) and held-out test (*n* = 75, 318) sets. Every record from one participant remained in one split. Of these participants, 351,166, 75,620 and 75,306, respectively, had at least one usable time-indexed record and contributed to world-model fitting or evaluation. We estimated preprocessing statistics and model parameters in the training set, selected checkpoints and calibration transforms in the validation set, and opened the test set only after the relevant analysis was fixed. External validation used CARDIA, ARIC and WHI, three independent US longitudinal cohorts, and the All of Us Research Program. Initial model training and checkpoint selection used UK Biobank; evaluation in CARDIA, ARIC and WHI adapted normalization parameters using pre-landmark histories, and All of Us was evaluated zero-shot, as described below. Modality-specific analyses included all eligible participants in the designated split; figure legends report the resulting sample sizes.

### Construction of time-resolved pan-modal records

We organized each participant’s data into permanent context and asynchronous observations. Permanent context comprised 40 genetic principal components, birth year, sex and ethnicity. Continuous variables were accompanied by missingness indicators; sex used female, male and unknown categories, and ethnicity used 20 reported categories plus an unknown category. This context remained available throughout a participant’s trajectory.

The dynamic record contained 1,257 clinical-event identities, 3,620 numeric features (374 laboratory features, 62 physical measures, 12 lifestyle features, 2,923 plasma proteins and 249 metabolites) and two volumetric imaging modalities, T1-weighted brain MRI and body MRI. Thus, 4,877 event or feature identities and two image types could revise the latent state when observed. For brain MRI, we reoriented the defaced T1-weighted image to canonical orientation, resampled it to 2-mm isotropic voxels, cropped or zero-padded it to 160×160×128 voxels around the centre of the foreground and linearly rescaled foreground intensities between the 0.5th and

99.5th percentiles to [0, 1]. For body MRI, we downsampled each intensity-standardized volume by a factor of 0.575 with linear interpolation, centre-cropped or zero-padded it to 128 × 96 × 208 voxels and clipped intensities to [−5, 5]. At each proteomic visit, all Olink measurements were represented jointly as one timestamped observation vector; repeated visits were retained as separate observations.

We expressed time as attained age in days. When an assessment date was available, we calculated visit age from the event-stream baseline age and the exact date difference. When only integer age was available, we used the conservative upper edge of the reported year; observations without a defensible time were excluded. Repeated visits remained separate. We combined numeric modalities recorded at the same attained age into one observation node. Clinical events at that age were processed first because their order within the day was not identifiable. No artificial no-event landmarks were inserted during world-model training, and no observation recorded after a query time was exposed to the model.

For each numeric feature *j*, we estimated the training-set mean *μ _j_* and standard deviation *σ_j_* and computed

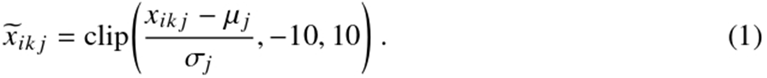

We preserved missing values and supplied a finite-value mask to every numeric modality encoder. The model therefore distinguished a missing measurement from an observed value near the training mean.

### Continuous-time latent-state model

HealthFlux used one continuous-time recurrent state-space model to represent both progression between observations and revision when new evidence arrived (Fig. 1b). Between observations, a neural ODE evolved the deterministic state; at each observation, a stochastic gate formed a prior and a posterior for the stochastic state, and a gated recurrent unit revised the deterministic state. Both steps ran in one recurrent loop and were optimized jointly. An encoder mapped the permanent variables and their masks to a 128-dimensional context ***c****_i_*. A separate attained-age encoder transformed normalized age, squared age, log age and six sinusoidal frequency pairs into a 64-dimensional vector ***a***(*t*). The model initialized a 256-dimensional deterministic state from context and age at the first record.

Each EHR token had a 256-dimensional embedding. Each numeric modality had its own multilayer encoder, which received standardized values and an observation mask and returned a 256-dimensional vector. Brain MRI and body MRI each had a separate three-dimensional convolutional encoder with the PlainConvUNet encoder architecture of nnU-Net:^42^ six stages of two 3×3×3 convolutions with instance normalization and leaky ReLU activation, 32 to 320 channels and stride-2 downsampling at the start of stages two to six. Both encoders were initialized from the same publicly released Triad weights, which were pretrained with a masked-autoencoder objective on 131,170 three-dimensional brain, breast and prostate MRI volumes,^43^ and kept separate parameters. A spatial-pyramid readout average-and max-pooled the third-and final-stage feature maps to 3 × 3 × 3 and 2 × 2 × 2 grids, layer-normalized each pooled vector, projected them to 64 and 192 dimensions and concatenated the projections into a 256-dimensional MRI vector. We added a learned modality embedding to every available numeric or MRI vector and used softmax attention to pool the numeric and imaging modalities present in the same observation row into a permutation-invariant embedding ***o****_ik_*. EHR tokens were encoded as separate event rows. When EHR and other observations had the same attained age, the EHR rows preceded the pooled observation row; EHR tokens and modality vectors were therefore not pooled into one simultaneous node.

Between consecutive observation ages *t_i_*_,*k*−1_ and *t_ik_*, an age-and context-conditioned neural ordinary differential equation evolved the deterministic state:

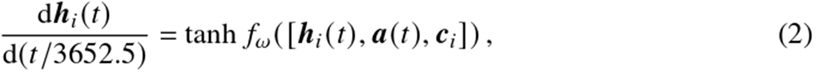

where *f_ω_* was a multilayer perceptron with two hidden SiLU layers. We used fixed-step fourth-order Runge–Kutta integration, a maximum step of 3,652.5 days and at most four substeps per interval. Explicit age conditioning allowed equal time intervals at different life stages to induce different state changes.

At each real observation, the evolved state 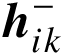 defined a 128-dimensional diagonal-Gaussian transition prior, and the new observation defined a posterior:

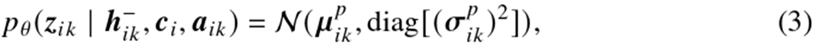

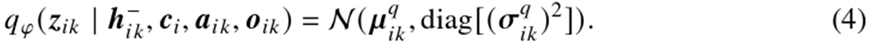

We sampled from the posterior during training and used its mean at evaluation. A gated recurrent unit then performed the observation jump,

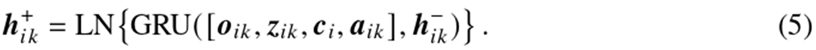

The model left the deterministic state at its ODE-evolved value when no observation was present. The final latent state was the layer-normalized concatenation

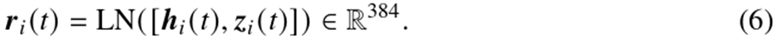

At observed nodes, ***r****_i_* (*t*) used the posterior mean. At an event-free query, the ODE evolved the previous state to the requested age and the transition prior supplied the stochastic mean. This construction produced a state at any requested time without implying that an unobserved assay or image had been measured. The action branch in Fig. 1b adds the effect of starting a medication to this state while every world-model parameter stays fixed; its drug encoder and state shift are described with the medication analyses below.

### World-model objective and optimization

We trained the latent-state model to predict the identity and timing of the next clinical event. From every eligible state, a categorical head predicted the next EHR token and a scalar head predicted log(1 + Δ*t*/365.25), where Δ*t* was the interval in days. The objective was

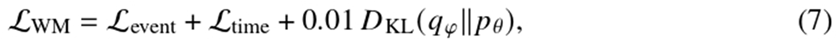

where L_event_ was next-event cross-entropy and L_time_ was the Smooth L1 loss for the transformed interval. Numeric observations updated the state but were not themselves next-event targets. Incident-disease labels and the downstream Health State Index were not used to fit the world model.

We initialized the EHR token-embedding table from a separately trained clinical-event model and both MRI encoder backbones from the pretrained Triad weights; we initialized all continuous dynamics, the remaining multimodal encoders, the MRI readouts and observation-jump parameters anew. All parameters, including both MRI backbones, were optimized jointly end to end, and no layer was frozen. Training used 351,166 participants and one randomly selected contiguous window of at most 96 observation nodes per participant per epoch. We optimized the model for 2,000 steps on six GPUs with a global batch size of 3,072, AdamW,^44^ an initial learning rate of 6 × 10^−4^, weight decay of 0.01, dropout of 0.1 and gradient-norm clipping at 1. The learning rate warmed up for 100 steps and then followed cosine decay to 10% of its initial value. We used bfloat16 mixed precision and accumulated the ODE solution in 32-bit floating point. Full validation was run every 50 steps, and we retained the checkpoint with the lowest total validation loss.

We audited temporal isolation in three ways. Encoding a fixed history was invariant to changes made after its endpoint; perturbing a future multimodal observation left all preceding states unchanged; and shifting attained age while holding the remaining inputs fixed changed the inferred state. The maximum numerical discrepancy in the two leakage tests was zero. These audits tested implementation-level causal masking and did not establish causal effects of exposures or diagnoses.

For the architecture ablations in Figure S1, we retrained two limiting cases with the same data, objective, EHR-embedding initialization, random seed and optimization budget (2,000 steps; effective batch size 3,072). Without ODE propagation, the deterministic state was carried unchanged between observations; without recurrent revision, the gated recurrent update was removed, so that observations informed only the instantaneous stochastic posterior. Each world model received a newly fitted five-year decoder with the primary architecture. All variants were evaluated on the same held-out participants by next-event cross-entropy, top-5 accuracy and absolute timing error, by timing error stratified by the preceding observation-free interval, and by five-year AUROC across the 1,256 diseases, averaged over those evaluable for every model, compared with 5,000 paired endpoint-bootstrap resamples. For the capacity ablation in Figure S2, the width of the internal multilayer perceptrons was set to 64, 96, 128, 192, 256, 512, 768 or 1,024 units (384 in the primary model) while the 384-dimensional health state was kept fixed; each model was trained and evaluated with the same protocol.

### Frozen-state disease readouts and Health State Index

We froze the world model before fitting the primary disease readout. The decoder received only the 384-dimensional state at one event-aligned landmark per participant and estimated five-year risk for 1,256 diseases and death. The landmark was the midpoint distinct clinical day, capped before the final clinical day; its state included every record on that day. Participants with the queried disease at or before the landmark were excluded from that endpoint’s risk set. The decoder standardized states using training participants and used a 384 → 1, 024 → 512 → 1, 257 multilayer perceptron with GELU activations, layer normalization and dropout of 0.08. We fitted the disease decoder in 286,651 training landmarks using endpoint-balanced binary cross-entropy. For endpoint *d*, the positive weight was 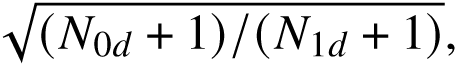 clipped to [1, 20]; output biases were initialized from training prevalence. AdamW used a learning rate of 2 × 10^−4^, weight decay of 10^−4^ and batches of 4,096. We trained for 18 epochs and selected the checkpoint with the highest validation mean AUROC. The primary five-year scores were not post-hoc calibrated, and no test observation contributed to fitting or selection.

For the single time-resolved rescue analysis in Fig. 5f, an auxiliary joint readout predicted conditional hazards for years 1–10 from the same frozen HealthFlux states. Cumulative products converted these hazards to monotone ten-year risks, and validation data selected the readout and estimated endpoint-and year-specific logit offsets. This auxiliary readout was retained only for the stated trajectory analysis; the uncalibrated fixed five-year decoder was the primary disease decoder and defined every primary HealthFlux comparison.

We derived the Health State Index (HSI) from a separate 384 → 64 → 16 → 1 readout while keeping the world model frozen. Survival, multimorbidity, incident-diagnosis count, current burden and future burden over 1, 3, 5 and 10 years jointly supervised the scalar, with ranking constraints orienting healthier outcome profiles above single-disease, multimorbid and terminal profiles. Each participant contributed unit total weight across repeated landmarks. We fitted the head in the training split and selected it by validation loss. One validation-derived population transform was then applied to every participant and time point: the 95th percentile of post-death raw scores defined the lower anchor, and the 95th percentile of first-observation scores among participants alive and free of all 195 modelled diseases for ten observed years defined the upper anchor. Raw scores were linearly mapped between these anchors to [0, 1] and clipped, and states after recorded death were set to zero. We never normalized a trajectory to its own baseline. HSI is therefore a common-reference prognostic index, not a percentage of biological health or a clinical decision threshold.

For Fig. 1g, we retained all 8,740 held-out participants who later met the prespecified multimorbidity definition and all 270,441 supported HSI states between ages 50 and 85. Background trajectories were coloured by participant mean HSI. Gaussian smoothing followed by shape-preserving piecewise-cubic interpolation was used only to render the dense background; highlighted trajectories used shape-preserving interpolation alone. Every curve was restricted to its observed model-support interval, with no backward or forward extrapolation. The highlighted participant identities had been fixed in the preceding case-selection analysis rather than selected using their final HSI values.

The HSI analyses in Figures S3 and S4 used the same world model and HSI head as Fig. 1g, without refitting, in held-out participants. The baseline HSI was the state after every record on the day of the first UK Biobank assessment (75,221 participants), and follow-up ended at death or at the end of administrative follow-up, as for Fig. 2h. The age reference chart sampled each participant’s HSI at every birthday between the first and last record from the most recent observation node (1.9 million person-years); centiles were smoothed over three years for display. Survival was summarized by Kaplan–Meier curves by baseline-HSI quintile, hazard ratios per 1-SD lower HSI came from Cox models adjusted for age and sex, and Harrell’s C for 10-year death was compared between age and sex with and without the HSI using 200 participant bootstraps. Ten-year cumulative incidence by baseline-HSI decile used the Aalen–Johansen estimator with death as a competing event, after excluding participants with the outcome at baseline. Multimorbidity was the first record of a second of 31 prespecified chronic-condition families; cardiovascular disease comprised I20–I25, I50, I60–I64 and G45, chronic obstructive pulmonary disease J40–J44, dementia F00, F01, F03 and G30, and cancer C00–C97 except C44. Self-rated overall health (UK Biobank field 2178) was recorded at the baseline assessment and was not a model input; differences from excellent health were adjusted for age, age squared and sex, with 2,000 participant bootstraps. For Figure S4, the HSI of every held-out participant was queried each quarter of age between the first and last record, as in Fig. 1g, so that each value used only records up to that time. For each of the 31 chronic-condition families, cases were participants whose first record fell at least two years after their baseline assessment. Each case was matched to up to five controls of the same sex whose baseline age differed by no more than six months and who were alive, under follow-up and free of the condition at the case’s age at first record; the HSI was not used for matching. Differences two years before the first record were averaged over cases with 2,000 case bootstraps, and mean trajectories used each case and its controls only at times at which both had a value, with 1,000 case bootstraps. When the same procedure was applied to every diagnosis code with at least 100 cases, 386 of 438 codes were preceded by a lower HSI; because several of the largest differences involved nonspecific codes recorded during health-care contact, Figure S4 shows the prespecified conditions.

### Out-of-distribution evaluations

The transfer experiments in Fig. 3 used HealthFlux states and semantic readouts fitted for each transfer setting. For the strict analysis of 1,256 diseases in panel b, HealthFlux was retrained independently for each exclusion fold under the protocol described below. Within every transfer comparison, participant splits, endpoint definitions and exclusion rules were fixed and shared across conditions.

We represented each disease or measurement identity by a fixed 1,024-dimensional text embedding generated with text-embedding-3-large. We L2-normalized the embeddings and projected them to 128 dimensions by uncentred singular-value decomposition fitted only to identities available for decoder training. The projection basis never used a held-out identity. These semantic queries allowed one shared readout to address identities without a trainable output row for each target.

For disease transfer, a bilinear decoder mapped a standardized latent state and disease query ***e****_d_* to

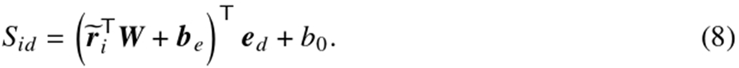

We trained ***W*** and the shared biases with balanced case–control logistic loss plus a pairwise ranking loss of weight 0.2. Each iteration sampled 64 disease queries, balanced across ICD-10 chapters, and 16 cases and 16 controls per query. We used AdamW for 2,000 iterations and selected the lowest validation objective. Wrong same-chapter descriptions and the mean description served as semantic controls.

We distinguished decoder-unseen disease from strictly unseen disease. In the decoder-unseen experiment, each held-out disease fold was absent from semantic-decoder fitting and from the singular-vector basis, while the common world model could have encountered that diagnosis. In the strict experiment, we divided the 1,256 diseases into five disjoint chapter-stratified folds, balancing training-case counts without using test outcomes. For each fold, we removed its diagnosis tokens from world-model inputs, next-event targets and the event-softmax support, reconnected the next eligible target after removal and trained the world model from random initialization without the transferred EHR embedding. The same fold was absent from semantic-decoder fitting and basis estimation. Every strict score therefore came from a model that had used neither the target diagnosis token nor its outcome labels.

For the whole-chapter comparison, we prespecified four ICD-10 chapters that contained at least ten evaluated endpoints with adequate incident cases and controls: circulatory (I00–I99; 22 endpoints), respiratory (J00–J99; 18), digestive (K00–K99; 27) and musculoskeletal (M00–M99; 27). Each chapter was withheld in turn. HealthFlux decoder-unseen models retained the common world model but excluded all target-chapter outcomes from decoder fitting and validation selection; strict models additionally excluded every chapter diagnosis token from world-model inputs, next-event targets and output support. We retrained RisQ from random initialization for each chapter while setting all target-chapter prognosis columns to its native ignore value in both training and validation; diagnosis history remained available, defining the matched decoder-unseen condition. No target-chapter labels informed checkpoint selection. All models were evaluated in the same prespecified held-out cohort of 61,504 participants and identical endpoint-specific five-year risk sets. Cohort eligibility required at least three distinct clinical-event days and support for a common event-aligned landmark and event-free query. We calculated AUROC and average precision for each endpoint, averaged endpoints equally within chapter and obtained 95% confidence intervals by 5,000 bootstrap resamples of endpoints.

For decoder-unseen measurement, we withheld eight laboratory and eight proteomic identities from readout fitting; the frozen world model could have encountered their values during representation learning. The training bank contained 53 laboratory and 128 proteomic identities with repeat observations. Given the frozen state at the source visit, the current standardized value, elapsed time, modality indicator and a semantic identity query, a shared decoder predicted the next observed standardized value. It combined a 128-dimensional state projection, the query, their elementwise product and six scalar time/value features. We fitted it with Smooth L1 loss for 3,000 iterations and selected the lowest tuning-set mean-squared error. Table 1b and Fig. 3c report the eight withheld laboratory identities, comprising 23,780 held-out repeat-measurement pairs, with pooled Pearson *r* = 0.629 and mean-squared error 0.574, versus 0.679 for carry-forward. Wrong-description, blank-description and carry-forward predictions served as controls.

For decoder-unseen population, the frozen world model had seen every demographic group, but the disease readout received no labels from the target domain. We evaluated male-to-female and female-to-male transfer, held out each attained-age quintile in turn, and held out each BMI quartile in turn. BMI was the latest available value at or before the landmark. For each disease and source domain, we fitted a class-balanced ridge classifier (*α* = 10) to the frozen states and evaluated it in the disjoint target domain. We required at least 20 source cases, 10 target cases and 50 controls in each domain. Training controls were capped at 20,000 and at five controls per case, with a minimum of 2,000 when available. Within each held-out setting, eligible disease endpoints were weighted equally; settings were then weighted equally within the sex, age and BMI dimensions. This experiment measured transfer of a readout into a label-sparse group within UK Biobank; it did not test a world model trained without that population.

For external-cohort generalization in Fig. 3g, we applied the UK Biobank-trained HealthFlux world model and disease decoder to ARIC, CARDIA and WHI without outcome-supervised refitting. HealthFlux+TTT, Delphi-2M and RisQ were compared on the same common AUROC-estimable disease endpoints within each cohort. HealthFlux+TTT updated only world-model layer-normalization parameters using unlabelled pre-landmark next-event and event-time objectives; the disease decoder remained frozen, and post-landmark outcomes were not used. Delphi-2M and RisQ were evaluated without test-time adaptation. AUROC was calculated separately for each disease and summarized across endpoints within each cohort.

For zero-shot evaluation in the All of Us Research Program (Fig. 3f), we applied the UK Biobank-trained HealthFlux, Delphi-2M, MILTON, RisQ, XGBoost and LightGBM models without retraining or test-time adaptation. For each participant, the index date was the first eligible physical-measurement date, and only information recorded before that date was available. The data available in All of Us comprised diagnosis history, age, sex, clinical laboratory tests, physical measurements and proteomics; each model used the subset of these inputs that it accepts. Each model predicted whether each disease or death occurred during the following five years. AUROC and AUPRC were calculated separately for each outcome and averaged with equal weight over the 971 outcomes evaluable for all models; 95% confidence intervals for the mean AUROC came from bootstrap resampling of outcomes. For each comparator, we report the mean paired outcome-level difference in AUROC and the proportion of outcomes for which HealthFlux had the higher AUROC. A score based on age alone served as a reference. Because the fitted per-endpoint XGBoost and LightGBM models from the UK Biobank comparison had not been retained, both were refit in UK Biobank with the same features, hyperparameters and random seeds before transfer; the XGBoost refits reproduced the original UK Biobank test predictions exactly, and the LightGBM refits changed the mean UK Biobank AUROC by less than 0.001.

### Medication-conditioned trajectory simulation

Medication use was taken from the regular medications self-reported at UK Biobank assessment-centre interviews. A medication was considered started when it was first reported, and the start was placed midway between that interview and the previous one; dose, dispensing and adherence were not available. Starting a medication was represented as an action that adds a learned residual to the dynamics of the frozen world model from the start time onwards, so that medication-on and medication-off branches begin from the same pre-medication state and diverge thereafter. A shared residual network received a medication-specific action vector. The world model, a decoder for six biomarkers (LDL cholesterol, apolipoprotein B, systolic and diastolic blood pressure, glucose and HbA_1_*_c_*) and the all-disease decoder were frozen.

Actions were trained on training-set participants who started a medication between two interviews. The medication branch was fitted to the biomarkers measured at the interview at which the medication was first reported: LDL cholesterol and apolipoprotein B for statins, systolic and diastolic blood pressure for blood-pressure-lowering drugs, glucose and HbA_1_*_c_* for metformin, and all six biomarkers for other medications, with penalties on changes in non-target biomarkers and on action magnitude. For statins, blood-pressure-lowering drugs and metformin, the simulated change in disease risk was additionally calibrated to effects per unit biomarker change from trial meta-analyses. At 1, 3 and 5 years after the start, the simulated log cumulative-hazard ratio of each target disease was constrained to the simulated biomarker reduction multiplied by the corresponding log hazard ratio: 0.76 for chronic ischaemic heart disease (I25) and 0.85 for stroke (I63–I64) per 1 mmol/L lower LDL cholesterol;^45^ 0.87 for stroke and heart failure (I50) per 5 mmHg lower systolic blood pressure;^46^ and 0.69 for type 2 diabetes (E11) per 1.65 mmol/mol lower HbA_1_*_c_*, the reduction achieved with metformin in the Diabetes Prevention Program.^47^ Statins comprised lipid-lowering drugs other than ezetimibe; blood-pressure-lowering drugs comprised calcium-channel blockers, angiotensin-converting-enzyme inhibitors, angiotensin-receptor blockers, *β*-blockers, *α*-blockers and diuretics. Actions were first trained for 81 commonly used medications and then extended to 259 UK Biobank medication codes with at least 10, 2 and 2 new users in the training, validation and test sets; codes corresponding to one of the 81 medications inherited its action, and every code was fitted to the follow-up biomarkers of its own new users without additional disease targets. Models were trained with three random seeds, and estimates were averaged across seeds.

Along each branch, five-year risks from the all-disease decoder were converted to annual hazards at 0, 1, 3, 5 and 10 years and integrated to cumulative hazards. For the screen (Fig. 4f), each code was simulated in the held-out participants who started it, from their state at the preceding interview. For each disease not yet recorded, the effect of a code was the median, across these participants, of the 5-year cumulative-hazard ratio between branches; a disease was counted when this ratio was 0.95 or lower, and the median across 1,256 diseases summarized the overall effect. For Fig. 4g, every code was applied to a shared reference cohort of 512 randomly selected held-out participants free of 21 prespecified major diseases at their first assessment: type 2 diabetes, ischaemic heart disease (I20–I25), heart failure, stroke and intracranial haemorrhage (I60–I64), chronic kidney disease, chronic obstructive pulmonary disease, dementia (F00, F01, F03, G30), breast cancer and prostate cancer. Ten-year risks of first major disease and of death were obtained from a five-year major-disease decoder trained on frozen HealthFlux states and from the death output of the all-disease decoder, and relative risk reduction was calculated as 100 × (*p̄*_off_ − *p̄*_on_)/*p̄*_off_, where *p̄* is the mean risk in the cohort. Candidate medications in Table 2 were restricted to codes with at least 30 held-out new users. Disease-system profiles (Figure S8a) were the median, across participants, of each participant’s median cumulative-hazard ratio within 14 groups of ICD-10 chapters, and reproducibility was assessed by comparing estimates between the three independently trained seeds (Figure S8b,c). The trajectories in Fig. 4a were simulated for statin and antihypertensive initiation in the same way.

### Drug representations and clinical-trial evaluation

To evaluate medication effects across many drugs and trial populations, a second medication module represented each drug by its pharmacological content rather than by a medication-specific action. Both modules act on the same frozen world model and decoders and are trained on medication starts in UK Biobank; they differ in how a medication is represented and in where it enters the model. Medications recorded at UK Biobank assessment-centre interviews were mapped to active ingredients with a published crosswalk,^48^ and the 438 ingredients reported by at least 100 participants formed the medication vocabulary. Each ingredient was described by its ATC codes, its ChEMBL molecular targets and mechanisms of action,^49^ an ECFP4 structural fingerprint^50^ and a BiomedBERT embedding^51^ of its name and mechanism of action. A drug encoder fused these views into a 64-dimensional embedding *e*, to which a learned identity embedding was added; the identity embedding was dropped with probability 0.5 during training so that effects could also be inferred from the descriptive views. Starting a set of medications shifted the 384-dimensional HealthFlux state ℎ by

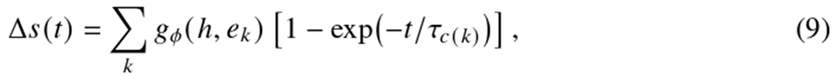

where *g_φ_* is a multilayer perceptron, *t* is the time since initiation and *τ_c_* is a learned onset time for each first-level ATC class *c*. The world model and the all-disease decoder were frozen. Fourteen biomarkers (LDL cholesterol, apolipoprotein B, HDL cholesterol, triglycerides, total cholesterol, glucose, HbA_1_*_c_*, C-reactive protein, creatinine, systolic and diastolic blood pressure, weight, body-mass index and FEV_1_) were read out from the state by frozen ridge regressions fitted to visit states of training participants, in units of the training-set standard deviation.

The action module was trained on 74,907 pairs of consecutive assessment-centre interviews at least six months apart; in 41,397 pairs, at least one ingredient was recorded for the first time at the later interview. Participants kept their world-model data split, and ℎ was the state at the later interview before its measurements were incorporated. The loss combined the binary cross-entropy of incident diseases recorded during the five years after the later interview (truncated at 30 June 2023), predicted by the frozen decoder with the shift evaluated one year after initiation; the squared error of the 14 biomarkers measured at the later interview, predicted with the shift evaluated at half the inter-visit interval and weighted by 0.2; and an *L*_2_ penalty on the shift. Following the active-comparator, new-user principle,^52^ a shared term for starting any new medication, conditioned on the state and the number of new ingredients, was learned jointly and omitted from simulations, so that simulated effects are specific to the drug rather than to medication initiation in general. We used AdamW^44^ (learning rate 10^−3^, weight decay 10^−4^, batch size 1,024, 30 epochs), selected the checkpoint with the lowest validation loss and averaged predictions over three random seeds.

For physiological responses (Fig. 4b), we extracted from the ClinicalTrials.gov results database^53^ randomized comparisons in which one arm received one or more vocabulary ingredients in addition to the regimen of the control arm (placebo or a shared background therapy) and that reported the mean or least-squares mean change from baseline in one of the 14 biomarkers after 4 to 104 weeks. Drug identities were taken from arm titles; comparisons involving active interventions outside the vocabulary and kinetic outcomes, such as clearance or synthesis rates, were excluded. Between-arm differences were converted to UK Biobank standard-deviation units, and differences of five or more standard deviations were excluded as unit errors. Contrasts that conflicted with established pharmacology, such as an increase in LDL cholesterol with a lipid-lowering drug, were checked against the source records, and four trials with misread comparator arms or units were excluded. The remaining 631 contrasts from 286 RCTs were pooled by inverse-variance weighting into 350 drug–biomarker pairs. Each trial was emulated in up to 1,500 held-out UK Biobank participants at their baseline visit who did not use the trial drugs and met the trial’s sex criterion and, when at least 300 participants remained eligible, its age range and a recorded diagnosis matching the trial condition. Both arms were simulated from the same states, and the simulated between-arm difference was read out at the trial duration, capped at two years. For an internal comparison (Figure S7a), we used held-out participants with two interviews: for each ingredient with at least 30 new users, the mean change in each biomarker between interviews among new users minus that among at least 100 non-users was compared with the simulated change at half the inter-visit interval, weighting pairs by 1/SE^2^. Sensitivity analyses (Figure S7c) took drug identities from arm descriptions rather than titles, restricted comparisons to placebo controls, or omitted the additional checks and source-record review. For disease and safety outcomes (Fig. 4c–e), binary outcomes were taken from ClinicalTrials.gov outcome-measure and adverse-event records and from primary trial publications and were mapped to the ICD-10 endpoints of the all-disease decoder or to death; aggregate categories, such as any serious adverse event, were excluded. Each comparison of a drug with its control for one outcome within a trial formed one observation, with the log odds ratio computed with a 0.5 continuity correction; reports of the same trial were linked into a single trial unit. The trial-effect model was a gradient-boosted regression-tree ensemble^54^ that predicted the log odds ratio from three groups of inputs. HealthFlux representations comprised the drug embedding *e* (without the identity component) and the endpoint’s output weights in the frozen all-disease decoder, each reduced to six principal components and averaged over ingredients or ICD-10 codes. Trial design comprised whether the outcome was reported as a clinical endpoint or as an adverse event, the log total sample size, the follow-up duration, whether the comparator was another drug, and the outcome group (coronary heart disease, stroke, heart failure, atrial fibrillation, diabetes, kidney disease, death or ICD-10 chapter). Evidence from other trials comprised inverse-variance pooled log odds ratios, with their standard errors and numbers of trials, for the same drug and outcome, the same drug across outcomes, the same first-level ATC class and outcome group, and the same outcome across drugs; these were computed from training trials only and always excluded the target trial. Observations were weighted by 1/(SE^2^ + *τ*^2^), where *τ*^2^ is a moment estimate of between-comparison heterogeneity. The model was fitted to drug-versus-control and head-to-head comparisons together and evaluated on comparisons of a drug with placebo or no added drug that had at least ten events.

Trials were the unit of all data splits. A random 20% of trials formed a sealed test set. In the remaining development trials, input groups and hyperparameters (maximum depth 3, learning rate 0.05, 150 iterations, minimum leaf size 60, *L*_2_ regularization 1) were selected by five-fold trial-grouped cross-validation. The sealed test set (46 RCTs, 105 comparisons) was evaluated once, with the model refitted on all development trials. Figure 4c,e and the deciles in Fig. 4d show ten-fold trial-grouped cross-validation over all trials with the selected configuration, so that each comparison was predicted by a model fitted without its trial; because the configuration was selected on development trials, these estimates may be slightly optimistic, and the sealed test set provides the unbiased estimate. As a reference, meta-analytic extrapolation predicted each comparison by the pooled estimate for the same drug and outcome in the training trials, or by no effect when none was available.

Agreement was summarized by the precision-weighted Pearson correlation *r_w_*, with weights 1/(SE^2^ + *τ*^2^), and by the proportion of comparisons with *P* < 0.05 in the trial whose predicted direction matched the observed direction. The 95% CIs came from 2,000 bootstrap resamples of drugs (Fig. 4b) or trials (Fig. 4c,d). The binned mean in Fig. 4b is the mean simulated change within eighths of the observed change. For calibration (Fig. 4d), comparisons were grouped by predicted value, and observed log odds ratios were pooled within each group with the same weights. Figure 4e shows the twelve comparisons with the largest absolute *z* statistic among those with *P* < 0.05, retaining one per drug–outcome pair.

### Clinical evaluation and statistical analysis

For the primary disease task, with *t_i_* denoting the landmark and *T_id_* the first record of disease *d*, we defined

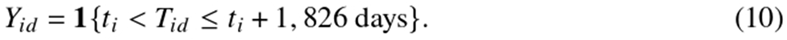

Participants without a recorded endpoint in this interval were treated as controls. Death did not censor other disease outcomes, and we did not model administrative censoring; estimates therefore refer to recorded incidence in the linked data. We quantified discrimination with AUROC and average precision across 1,256 diseases and death. An endpoint contributed to the mean only when its risk set contained both cases and controls. A duplicated O01 code was represented once, retaining the source token with more training cases. Endpoint definitions were fixed without reference to model predictions, and test outcomes were not used for fitting or checkpoint selection. Mean AUROC and mean average precision are the unweighted arithmetic averages of endpoint-specific values, with every estimable endpoint receiving equal weight. Model comparisons used the same 61,504 held-out landmarks and identical endpoint-specific risk sets for HealthFlux, Delphi-2M,^21^ RisQ,^23^ XGBoost,^34^ LightGBM^35^ and MILTON.^22^

XGBoost and LightGBM were fitted separately for each endpoint using the same causal 213-dimensional event-history representation and validation-only model selection. We used the official MILTON implementation (commit 430fb86) and its phenotype-specific 67-biomarker pipeline, fitting a separate model for each endpoint with the original preprocessing and case– control sampling. MILTON’s native five-fold hyperparameter search was used when both classes had at least five training examples; for rarer endpoints, for which five-fold selection is undefined, we retained its prespecified XGBoost configuration. This adaptation harmonized prediction landmarks, risk sets and five-year outcome definitions without adding EHR-sequence modelling to MILTON. Delphi-2M was evaluated from its frozen native checkpoint using each endpoint token’s log-rate. RisQ was fitted jointly to the prognosis queries with its official Rep-Query pipeline, selected only on the validation split and evaluated using its native five-year cumulative-max logit. All comparisons used native risk scores as ranking functions; no test-set calibration or model selection was performed.

For the age profiles in Fig. 2a, we formed 20 contiguous 2.5-year bins from ages 30 to 80. The four qualitative incidence archetypes were identified from smoothed training-set incidence and required qualitative replication in the validation split; the held-out test curves were not used for disease selection. Endpoint-specific logistic calibration slopes and intercepts were fitted in validation data. The figure shows unsmoothed test-set incidence with Wilson intervals and the mean calibrated HealthFlux prediction in each bin. This calibration was used only for the probability-profile display and did not alter the uncalibrated primary discrimination comparison. HealthFlux estimates the risk of 1,256 diseases and death. Disease-averaged metrics were computed over the diseases with at least one incident case and one control in the held-out risk set for every compared model; diseases without held-out cases cannot be evaluated and did not enter the averages. An AUROC is the probability that a randomly chosen future case is assigned a higher predicted risk than a randomly chosen participant who remains free of the disease; in the main text, we therefore also express it as the number of such case–non-case pairs out of 100 that the model ranked correctly. Figure 2c groups five-year metrics for 1,256 diseases and death by ICD-10 chapter. Pregnancy and childbirth (O00–O99; 61 endpoints), perinatal conditions (P00–P96; 3 endpoints) and congenital anomalies (Q00–Q99; 58 endpoints) were analysed as one group, labelled Pregnancy & congenital in Fig. 2c,g and Fig. 5a,b, because the perinatal chapter alone was too small to summarize. Several complementary panels used prespecified HealthFlux predictions. For Fig. 2d, we reduced diagnoses to 15 ICD-10 chapters, excluded participants with the target chapter already recorded at the index landmark and computed binary Phi correlations between each recorded source chapter and each five-year incident target chapter or death. Same-chapter cells were undefined because the source indicator was then constant. We separately followed four prespecified source-to-target sequences from 0.25 to 5 years after the first source diagnosis. These were E11 type 2 diabetes to N18 chronic kidney disease, I10 hypertension to I25 chronic ischaemic heart disease, I25 chronic ischaemic heart disease to I50 heart failure, and J44 chronic obstructive pulmonary disease to J18 pneumonia. Curves are descriptive cumulative recorded-diagnosis proportions; uncertainty came from 1,000 participant-level bootstrap samples.

Figure 2e used a common 180-day window in 66,121 participants with paired frozen states. We standardized state dimensions by training-set moments, calculated their root-mean-square displacement, annualized the result and stratified participants by the number of diagnoses newly recorded inside the same interval. This analysis did not align participants to the next clinical event and did not impose an event-time discontinuity. Figure S5 stratifies the 61,504-participant held-out landmark cohort by the number of additional diagnosis events recorded before the landmark after the first diagnosis (0, 1–2, 3–5, 6–9, 10–19 or ≥20). Within each stratum, we averaged predicted five-year risk and observed five-year incidence over each endpoint-specific risk set and ranked the 1,256 diseases and death. We summarized concordance with Spearman *ρ*; two-sided 95% intervals came from 10,000 endpoint bootstrap resamples stratified by the 15 ICD-10 chapters, with death retained once per resample. The endpoint-level paired ranks are coloured by ICD-10 chapter. This is a cross-sectional case-mix analysis rather than a within-person history-ablation experiment.

For the controlled information-blackout experiment in Fig. 2f, HealthFlux and Delphi-2M started from the same participant-specific real-event landmark. We then withheld every post-anchor EHR event and every post-anchor laboratory, omic, imaging, lifestyle and physical observation from both models and queried them one to five years later. At each query, the outcome was the first record of each endpoint during the following 1,826 days, using identical dynamic risk sets and labels for both models. The displayed set contained 191 endpoints with at least 20 cases and 100 controls at the ten-year stress-test query; this support rule used outcomes only and was fixed for all displayed years. HealthFlux simulated the latent state by ODE integration of the deterministic component and the transition-prior mean for the stochastic component, without observation updates, followed by its frozen five-year decoder; Delphi used its frozen native no-event token and endpoint log-rate. We compared equal-endpoint mean AUROC and ΔAUROC (HealthFlux minus Delphi), with 95% intervals from 5,000 paired endpoint bootstrap samples. Because Delphi supplies a native log-rate rather than a calibrated five-year probability, this experiment evaluates ranking discrimination, not probability calibration.

For high-risk enrichment in Fig. 2g, we analysed the 1,256 diseases and assigned them to the 15 disease systems used in panel c. Within each endpoint-specific held-out risk set, we ranked participants by the frozen HealthFlux five-year score after excluding prevalent target disease and selected the top 1% and top 10%. Incidence enrichment was observed five-year incidence in the selected group divided by incidence in the complete endpoint-specific at-risk population. We then averaged disease-level enrichment without weighting within each chapter. For each endpoint, we additionally calculated the proportion of all incident cases that occurred in the top-10% group and the ratio of five-year incidence in that group to that in the remaining 90% of the at-risk population, and summarized both by their medians across endpoints. Two-sided 95% intervals came from 10,000 bootstrap resamples of endpoints within chapter. Death was audited separately but excluded from this panel because it is not an ICD-10 disease chapter. These fixed-window ratios are incidence enrichment, not time-to-event hazard ratios.

For the one-to-twenty-year comparison in Fig. 2h, both models started from the same event-aligned landmark in the 61,504-participant held-out cohort, and no observed post-landmark event or measurement was supplied. The prespecified targets were the same 1,256 diseases and death used in the primary comparison. HealthFlux used the frozen world model. At each simulated step, its event head supplied validation-temperature-scaled next-diagnosis probabilities and its time head supplied a next-event interval augmented by a validation-derived residual; the ODE advanced the state to that time, the sampled diagnosis was injected through the observation-update pathway, and generation continued until twenty years or death. Delphi-2M^21^ used its frozen native competing-exponential autoregressive process, with each sampled event and interval appended before the next prediction. Both generative models used eight Monte Carlo trajectories per participant and were summarized by particle-averaged cumulative conditional-incidence scores. Participant and endpoint order were verified before evaluation.

At each annual horizon, cases had a first recorded endpoint by that horizon, controls remained endpoint-free and under observation through it, and prevalent endpoints were excluded participant-wise. Non-fatal endpoints were censored at death or the end of administrative follow-up, whichever occurred first; death was censored at the end of administrative follow-up. We calculated cumulative/dynamic AUROC with inverse-probability-of-censoring weights estimated within each endpoint risk set and truncated at 20. Curves are equal-endpoint means among diseases with estimable AUROC at each horizon; two-sided 95% intervals came from 10,000 endpoint bootstrap samples. These are ranking-discrimination estimates rather than calibrated absolute-risk probabilities.

For Fig. 2i, we screened 1,256 diseases using a validation selection subset (Figure S6a,b). The screen required at least 20 cases in that subset and 10 in a disjoint calibration subset, AUROC ≥ 0.70, sensitivity ≥ 0.20 at a 99%-specificity cutoff, and average precision at least twice the 90-day incidence. Thirty-five diseases met these validation criteria. Two had fewer than 30 cases in the held-out test set and were excluded for insufficient test support, leaving 33; test-set performance did not determine disease selection. In the held-out cohort, each disease’s first recorded diagnosis defined a diagnosis-aligned case episode. At 120, 90, 60, 30 and 10 days before that diagnosis, the frozen world model received only the record available by the query time and estimated the probability of a first diagnosis during the next corresponding 120, 90, 60, 30 or 10 days. The disease-and horizon-specific alarm cutoff was the 95th percentile of non-case risk scores at validation-set visit landmarks. The exploratory high-history-burden subgroup required at least four diagnoses during the year preceding day −120, assessed before inspecting test scores. The main panel shows the 32 candidates with at least ten such cases (5,494 unique participants; 10,304 participant–disease episodes); O70 had one high-burden case and was omitted from this display. Figure S6c shows all 33 diseases among 28,828 unique test participants and 62,820 participant–disease episodes, and Figure S6d compares the two case sets disease by disease. Each dot is the proportion of cases above its fixed cutoff; the median column summarizes disease-specific sensitivities with their interquartile range, not a pooled participant-level proportion. Since the cutoffs were selected at visit landmarks and the displayed cases were sampled relative to diagnosis, their local specificity and positive predictive value cannot be inferred from this plot. Figure S6e reports case detection and the separate fixed-visit predictive-value analysis at 95%, 97.5% and 99% validation-specificity settings.

The participant was the independent unit for participant-level analyses, and the disease was the unit for endpoint-mean summaries. We used paired participant bootstrap samples for matched predictions and disease bootstrap samples for comparisons of endpoint means. Unless stated otherwise, confidence intervals were two-sided 95% intervals. Wilson intervals described observed proportions. We controlled stated families of multiple comparisons with the Benjamini– Hochberg procedure.^55^ Tests were two-sided unless a directional alternative had been fixed in advance. This retrospective study involved no treatment assignment, randomization or investigator blinding.

### Disease-risk geometry and pan-modal interpretation

For the participant-level semantic projection in Fig. 3a, each of 75,619 validation participants was represented by the risk-weighted mean of the 196 fixed disease-description vectors. Principal-component analysis followed by UMAP^56^ produced the two-dimensional display. ICD-10 chapter, sex and the disease-associated age quintile were added only after projection. For the strictly unseen endpoints in panel b, transferred-head distance was one minus cosine similarity between the transferred and supervised disease-risk directions. We related distance to AUROC with Pearson correlation and an ordinary least-squares line; its 95% band was obtained from 1,000 endpoint bootstrap samples.

The modality-integration analyses in Fig. 5 used frozen HealthFlux states and prespecified disease readouts. Panels a–c used the primary five-year decoder in the 61,504-participant held-out landmark cohort, whereas panels d and e used their prespecified task-specific predictions. For panel a, each of the 1,256 diseases and death was represented by its 256-dimensional EHR event embedding from the frozen world model. These embeddings were learned from longitudinal event histories using the world-model objectives. We projected them with label-free cosine t-distributed stochastic-neighbour embedding (perplexity 75; 2,500 iterations; fixed seed). ICD-10 labels and density contours were added post hoc and did not influence coordinates. The two-dimensional axes and distances are descriptive.

We quantified modality use in the same frozen HealthFlux model with complementary gradient and ablation analyses.^57, 58^ We placed a scalar gate *g_m_* on modality *m*, multiplying standardized values and availability masks for numeric modalities and the input volume for MRI, and defined its absolute local attribution for disease score *S_id_* as

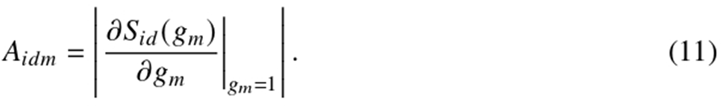

We averaged attributions among held-out participants in whom the relevant modality was observed. Displayed shares were normalized across nine biological modalities, excluding clinical records and demographic variables. Each map shows a disease’s attribution share minus that modality’s mean across 1,256 diseases and death. Chapter enrichment compared a chapter mean with the corresponding full-panel mean; global Benjamini–Hochberg FDR controlled the complete chapter-by-modality family, and leave-one-modality-out sign agreement provided a robustness flag. In panel b, point area represents the absolute chapter-to-global attribution difference. Dark rims mark global Benjamini–Hochberg FDR < 0.10; gold rims additionally require leave-one-modality-out sign concordance. For panel d, which used the prespecified multi-horizon readout, we separately masked the indicated input values and availability indicators, including EHR history and genetics, or removed the MRI observations, while holding every parameter fixed. Root-mean-square score change was computed over 196 endpoints and the one-, five-and ten-year horizons, and uncertainty came from participant bootstrap samples. These analyses describe use by the fitted model and do not establish biological importance or a causal effect. We retained 3,567 measured features that were observed in at least 100 held-out participants and had non-degenerate fingerprints: 40 genetic principal components, 180 blood-chemistry features, 102 haematology features, 62 physical-function traits, 12 lifestyle indicators, 2,922 proteins and 249 metabolites. MRI, which entered the model as images, was analysed at the modality level in panels a, b and d. Each feature was represented by its signed disease-response fingerprint across the 1,257 five-year outputs, obtained with a feature-level gate defined as above but without the absolute value and averaged over participants in whom the feature was observed, and Pearson correlations quantified similarity between fingerprints. The network displayed every retained feature. For each node, we selected the two strongest positive and one strongest negative within-modality correlations and the two strongest positive and one strongest negative cross-modality correlations, retained links with |*r* | ≥ 0.60 and merged duplicate undirected links. This produced 15,044 display edges. A seeded spring layout arranged modality communities by positive cross-modality connectivity; two-component randomized principal-component analysis of row-centred, unit-normalized fingerprints placed features within each community. Edge selection and layout provided a reproducible visual backbone rather than a significance-thresholded physical interaction map. Aggregating input-sensitivity patterns into feature representations follows the general principle of attribution-based model embedding.^59^

We constructed protein-and metabolite-derived disease geometries by correlating molecular association profiles between diseases. Their agreement was evaluated across all 19,110 unordered disease pairs by Pearson correlation and ordinary least-squares fitting, with disease-label permutation and within-chapter analyses as sensitivities.

The hypertension rescue panel combined frozen single-modal comparator outputs with panmodal HealthFlux outputs. The parent cohort for each modality comprised participants whose first I10 record occurred between ages 65 and 75 and whose age-65 score fell below that single-modal comparator’s threshold at nominal 90% specificity in participant-disjoint controls. The pan-modal score was not used to form this miss cohort. We applied the independently validation-fixed pan-modal HealthFlux threshold, also at nominal 90% specificity, and defined the rescue fraction as HealthFlux-positive cases divided by all true cases missed by the corresponding single-modal model. Ten-year HealthFlux risk came from the auxiliary annual-hazard readout described above; the single-modal probabilities and parent cohorts were not refitted. Displayed nodes were the exact age-65 query and genuine prediagnosis post-EHR-event states, with a one-day target guard and no fictitious events, interpolation or future extrapolation. Within each facet, the thick paired curves show the eligible participant with the greatest mean pan-modal-minus-single-modal gap among cases meeting the long-span display rule; thin curves show all rescued participants.

For Fig. 5g–i, cases with a future first record of L02 cutaneous abscess, J96 respiratory failure or G47 sleep disorder were compared with age-, sex-, visit-and diagnosis-history-matched controls at annual landmarks from ten to one years before diagnosis. At each landmark, the frozen one-year HealthFlux risk score and standardized biomarker values used only the causal record prefix. Biomarker gaps were oriented so that positive values denoted the adverse direction. Real visits only were used; we applied no carry-forward or monotonic post-processing, and unsupported years remained missing. Display curves used an *n*-weighted local-linear smoother with a 1.65-year bandwidth and required at least eight cases and eight controls in a supported bin. Paired participant bootstrap samples quantified risk-gap uncertainty. The displayed waist-circumference/grip-strength, aspartate-aminotransferase/glucose and C-reactive-protein/weight examples were selected for temporal concordance and are hypothesis-generating rather than causal.

### MRI analyses

We estimated cross-sectional age trajectories for tissue composition, regional volume and cortical morphology with restricted cubic splines adjusted for sex, imaging site and calendar time. Models of size-dependent traits additionally included intracranial volume. A separate Poisson model estimated the age-associated rate of diagnoses during follow-up. These curves provide population context and do not represent repeated within-person brain ageing.

The model-dependent MRI panels in Fig. 6b–f used HealthFlux with fixed participant splits and model parameters throughout.

We measured regional model dependence by replacing, in each participant’s model-input brain volume, the voxels of one of eight subcortical structures segmented with FIRST^60^ (left and right combined) with that structure’s mean input intensity in training participants, while holding all other inputs and disease readouts fixed. Analyses used training (*n* = 18, 216) and held-out test (*n* = 3, 933) participants with brain MRI; the state was read at the first brain scan, and scores were five-year decoder logits for 195 diseases in participants without a prior record of the disease. We estimated full-minus-ablated score changes separately in training and held-out participants and compared the 1,560 disease–region pairs. Replacement with zero intensity produced larger score changes with a broadly similar ordering of disease–region effects (held-out Spearman *ρ* = 0.73). Within each disease, we normalized root-mean-square regional effects, which also scale with structure size, and summarized brain-related, visceral or metabolic and other diseases using 20,000-replicate disease-bootstrap intervals.

For the case studies in Fig. 6e,f, we used each training or validation participant’s most recent scan at or before the fixed five-year prediction time; future cases and at-risk controls followed the five-year labels of the disease readouts. Model sensitivity was the eight-point path-average gradient of the frozen five-year disease logit with respect to the preprocessed MRI volume, along a straight path from an all-zero volume, averaged over at-risk training participants (393, 320 and a fixed-seed sample of 512 for chronic ischaemic heart disease, lipoprotein metabolism disorders and type 2 diabetes, with 3, 5 and 4 validation future cases, respectively). Brain gradients were returned to native space, mapped to the ICBM152 template with each participant’s UK Biobank T1-to-MNI affine transformation^36^ and averaged within an 8 × 8 × 8 grid. We retained 140 brain cells within the template brain mask with at least 20% voxel coverage in at least half of the training participants. Body gradients were averaged on the same grid in the model’s body-image space (512 cells). Observed morphology was the validation future-case-minus-control difference in grid-cell intensity of MNI-space T1 images (brain) or model input volumes (body). Cell intensities were adjusted for age, squared age, sex, scan year and, for brain MRI, brain-mask fraction with training-set regression coefficients; differences and model sensitivity were expressed in training residual standard deviations. We calculated spatial Spearman correlations across unsmoothed supported cells and obtained 95% intervals from 1,000 bootstrap resamples of validation cases and controls with model sensitivity held fixed. Display smoothing was support-normalized. The grid provides coarse spatial context and does not support voxelwise localization.

## Supplemental information

**Figure S1:**
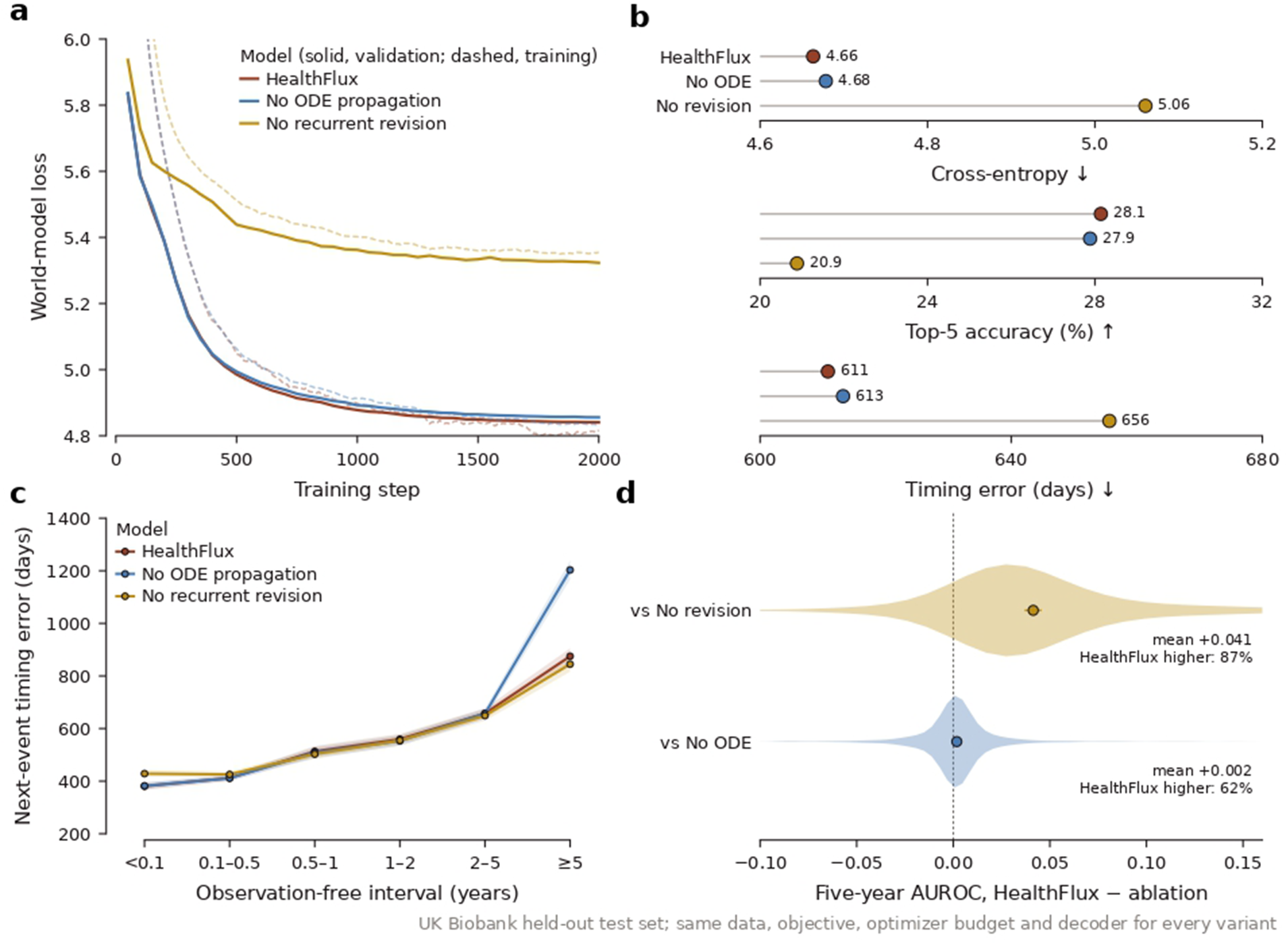
Architecture ablations of the HealthFlux world model. **a**, World-model loss during training (dashed, exponentially smoothed) and on the validation split (solid) for HealthFlux and two retrained temporal-pathway ablations: without ODE propagation between observations and without gated recurrent revision at observations. **b**, Held-out next-event cross-entropy, top-5 accuracy and absolute error in next-event timing. **c**, Next-event timing error by the length of the preceding observation-free interval for HealthFlux and both ablations; bands are 95% bootstrap intervals. **d**, Paired differences in five-year AUROC (HealthFlux minus ablation) across the 1,256 diseases (averaged over those evaluable for every model); violins show the endpoint distribution, and points and bars show means with 95% endpoint-bootstrap intervals. All variants used the same data, objective, embedding initialization, random seed, optimization budget and decoder architecture and were evaluated on the same held-out participants.

**Figure S2:**
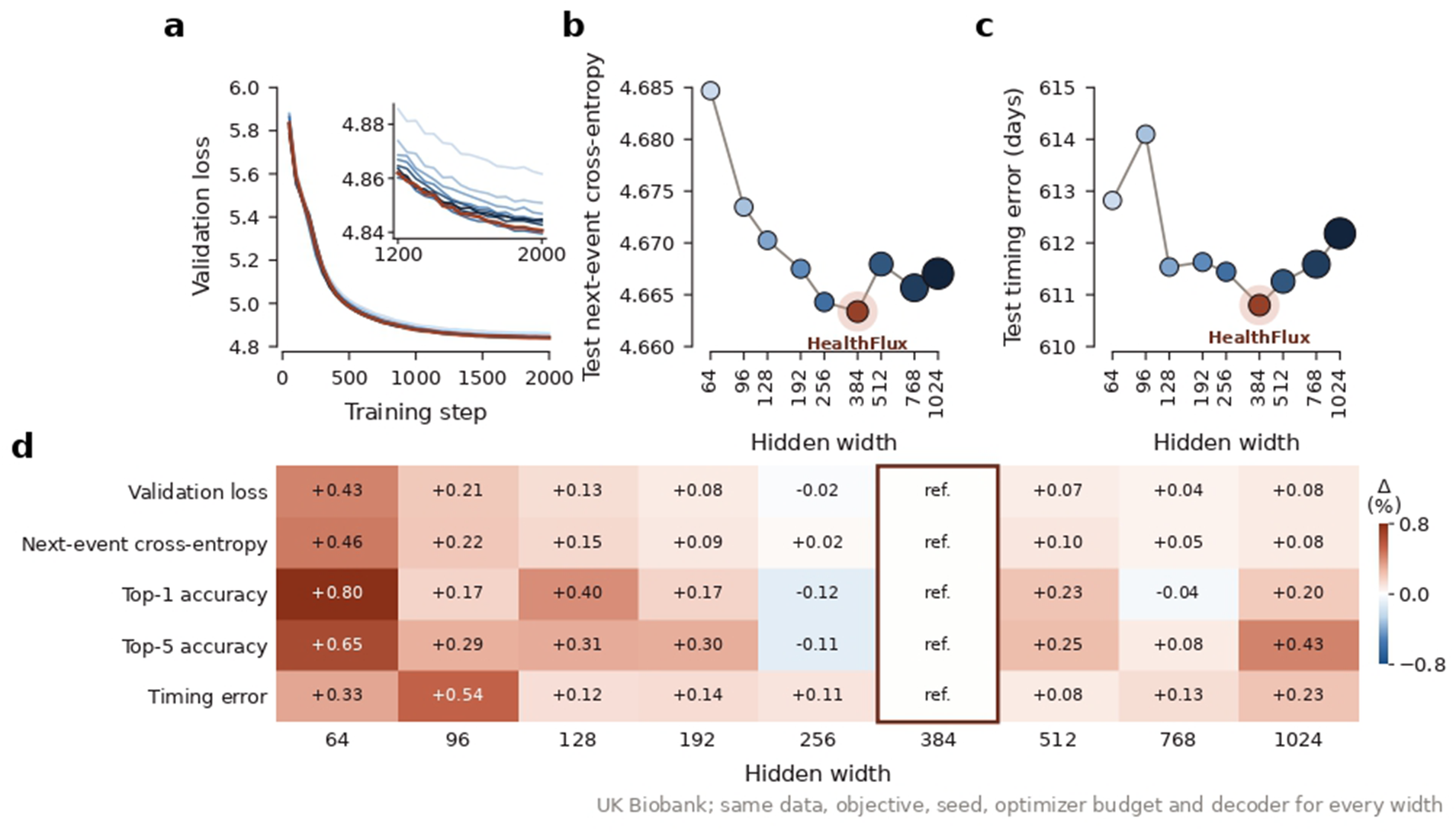
Model-capacity ablation of the HealthFlux world model. The width of the internal multilayer perceptrons was varied from 64 to 1,024 units (4.5 to 18.5 million parameters) while the 384-dimensional health state, data, objective, seed, optimization budget and decoder architecture were held fixed; width 384 is the primary HealthFlux model. **a**, Validation loss during training; the inset enlarges steps 1,200–2,000. Colours run from narrow (light blue) to wide (dark blue), with HealthFlux in red. **b,c**, Held-out next-event cross-entropy (**b**) and absolute timing error (**c**) by hidden width; marker area is proportional to the number of parameters. **d**, Relative difference from HealthFlux for each width and metric, oriented so that positive values indicate worse performance than HealthFlux.

**Figure S3:**
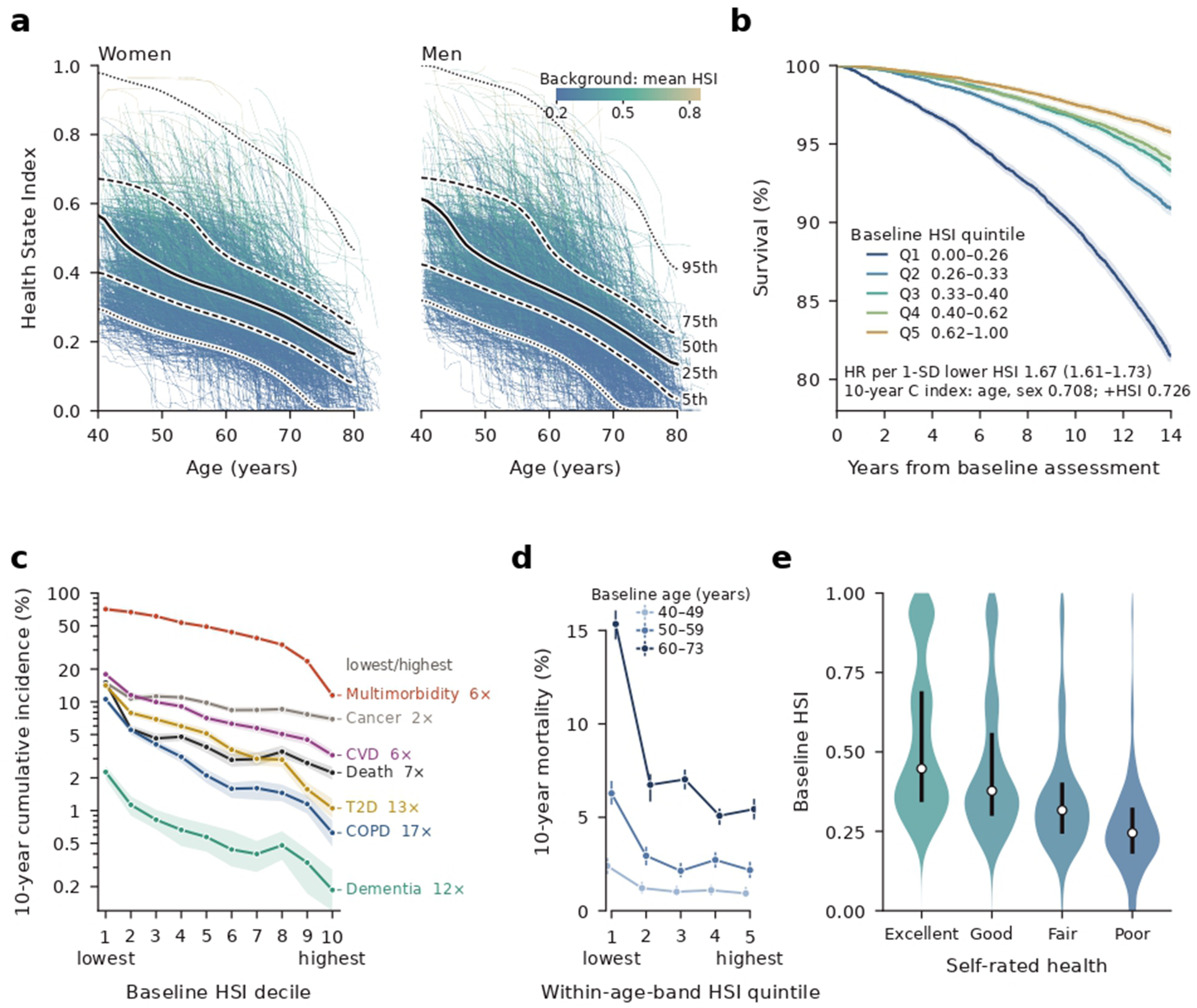
Population validity of the Health State Index. **a**, HSI by age in held-out women and men. Background, 1,800 trajectories per sex at recorded observations, coloured by participant mean HSI as in Fig. 1g; lines, 5th, 25th, 50th, 75th and 95th centiles of annual person-year samples. **b**, Survival by quintile of HSI at the baseline assessment (75,221 participants), with the age-and sex-adjusted hazard ratio per 1-SD lower HSI and the 10-year concordance for death. **c**, Ten-year cumulative incidence by baseline-HSI decile, with death as a competing event and prevalent cases excluded; labels give the ratio of incidence in the lowest to that in the highest decile. CVD, cardiovascular disease; T2D, type 2 diabetes; COPD, chronic obstructive pulmonary disease. **d**, Ten-year mortality by HSI quintile within three baseline age bands. **e**, Baseline HSI by self-rated overall health, which was not a model input (74,740 participants: excellent 12,136, good 43,316, fair 15,843, poor 3,445); white dots, medians; bars, interquartile ranges. In **b–d**, bands and bars denote 95% confidence intervals. The HSI head was trained on survival and disease burden in training participants; these results describe its performance in held-out participants.

**Figure S4:**
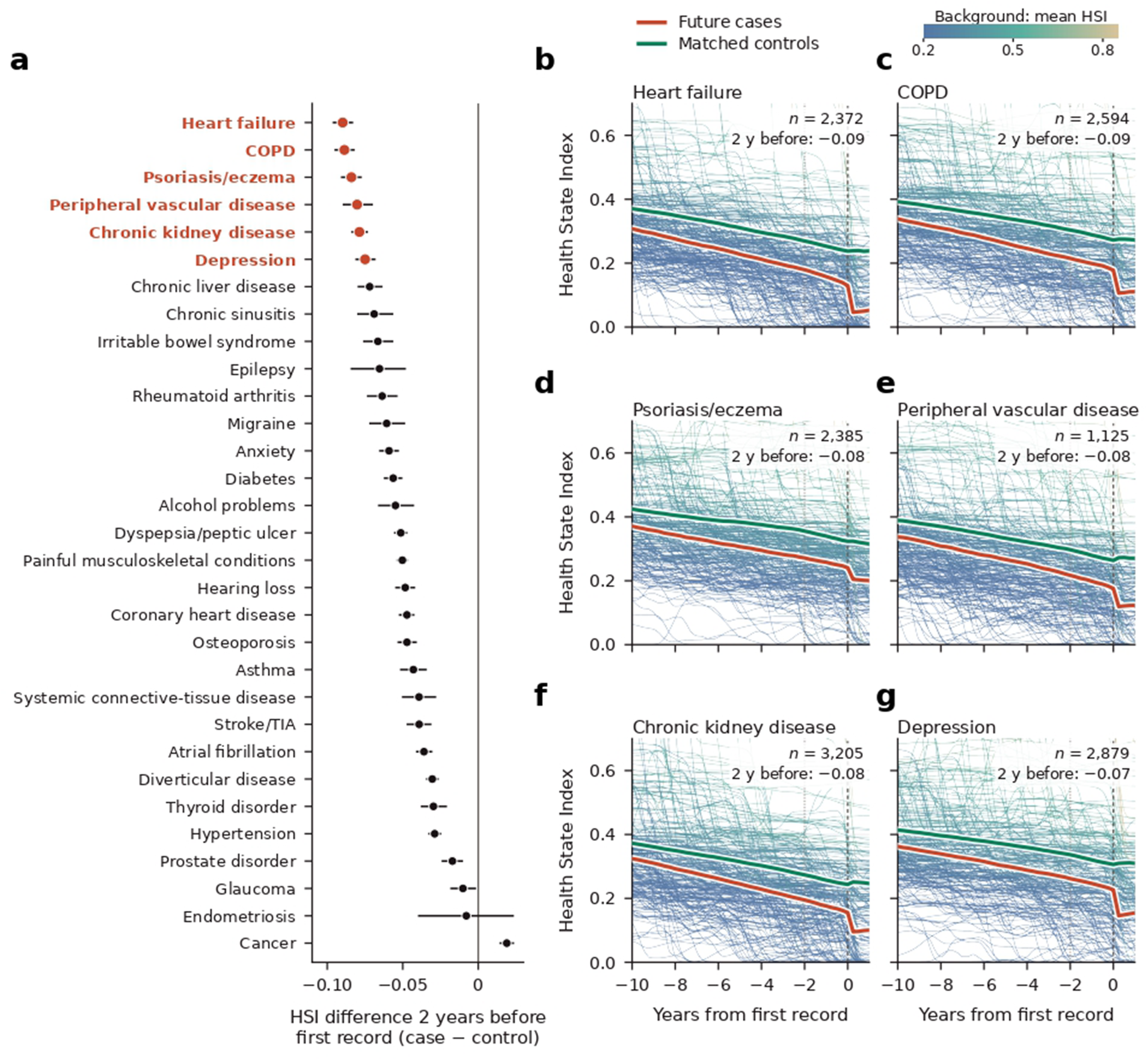
Health State Index before the first record of chronic conditions. **a**, Difference in HSI between future cases and matched controls two years before the first record of each of the 31 chronic conditions that define multimorbidity in Fig. 1g (196 to 11,865 cases per condition). Cases were first recorded at least two years after the baseline assessment, and each was matched to up to five controls of the same sex and baseline age who were free of the condition at that age. Points and bars, means and 95% confidence intervals; conditions in rust are shown in **b–g**. Cancer comprises non-melanoma skin, breast and prostate cancer, as in the multimorbidity definition. **b–g**, HSI from ten years before to one year after the first record of the six conditions with the largest differences, drawn as in Fig. 1g. Background, individual trajectories of 150 cases and 150 matched controls, coloured by participant mean HSI; lines and bands, means and 95% confidence intervals of future cases (rust) and matched controls (green); dotted line, two years before the first record. Numbers give cases and the difference at two years.

**Figure S5:**
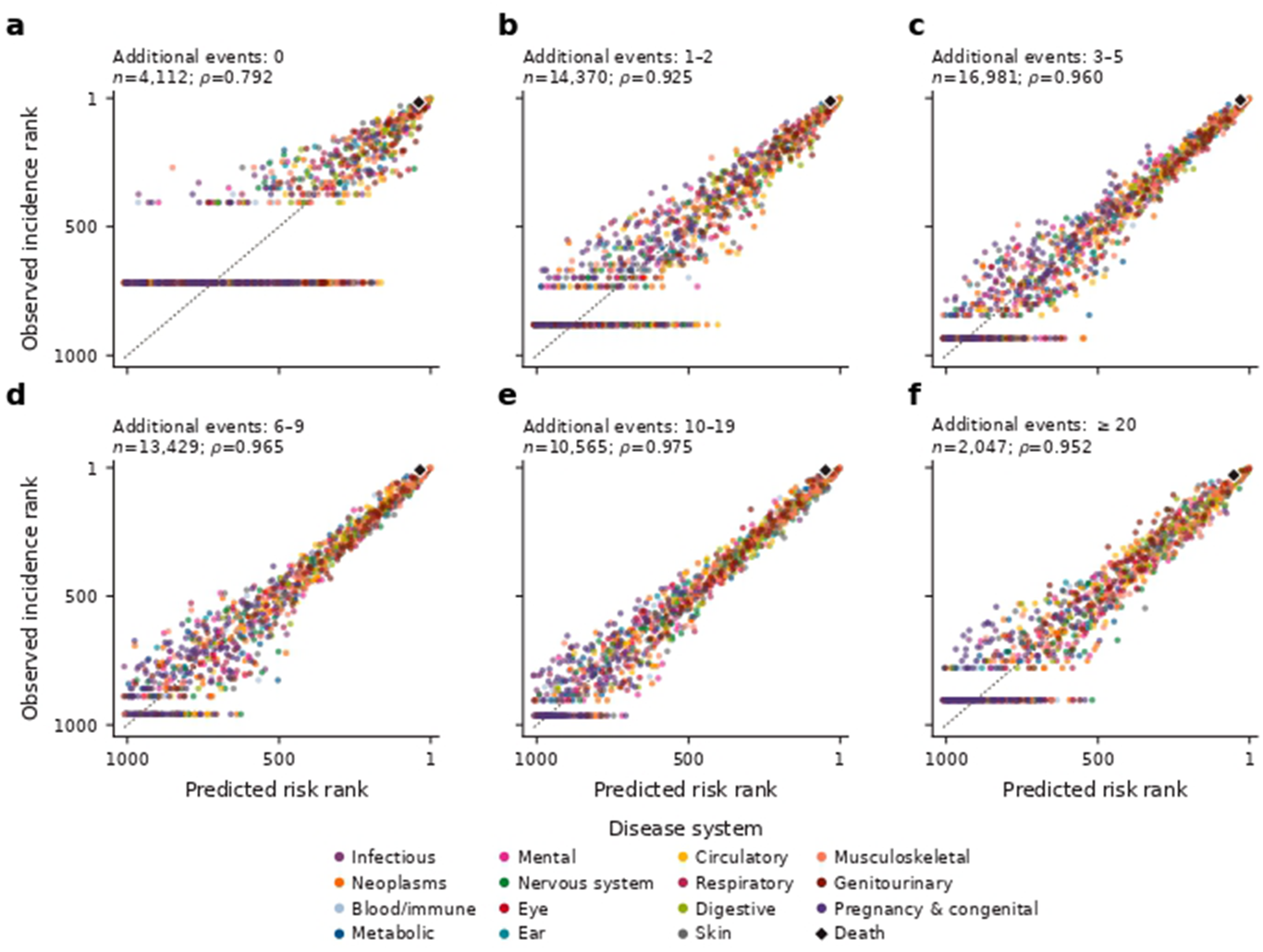
Disease-level rank concordance across recorded-diagnosis histories. **a–f**, Predicted versus observed five-year incidence ranks for 1,256 diseases and death, stratified by the number of additional diagnoses before the landmark. Each point is an endpoint, coloured by disease system as in Fig. 2c; *n* gives participants in each stratum. Spearman *ρ* summarizes endpoint-level agreement.

**Figure S6:**
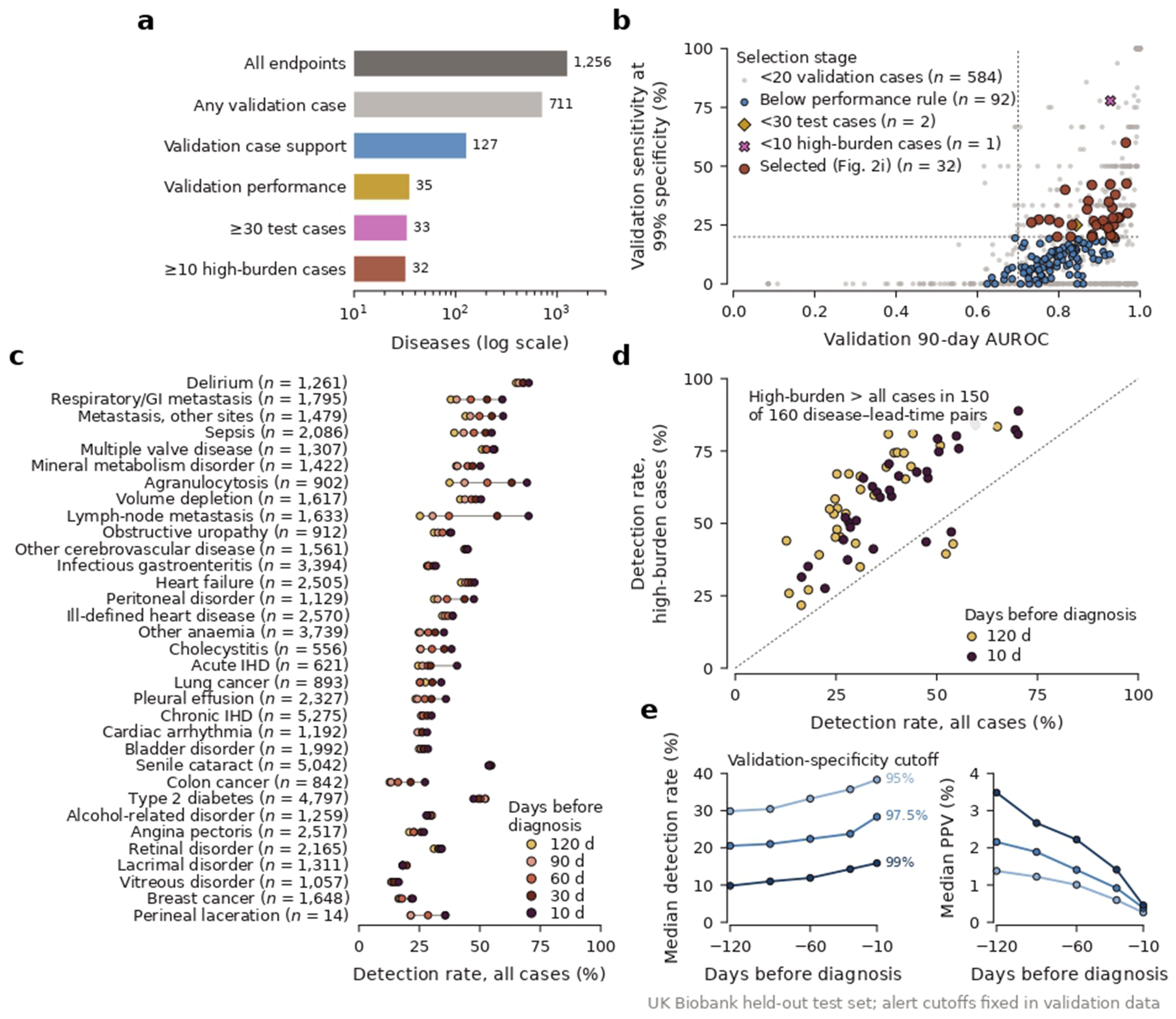
Selection of early-detectable diseases and prediagnostic detection across all cases. **a**, Selection path from 1,256 diseases to the 32 shown in Fig. 2i. Validation case support required at least 20 cases in the validation selection subset and 10 in a disjoint calibration subset; validation performance required 90-day AUROC ≥ 0.70, sensitivity ≥ 0.20 at a cutoff targeting 99% validation specificity and average precision at least twice the 90-day incidence. Test-set case counts, but not test performance, defined the last two steps. **b**, Validation screen of the 711 diseases with at least one validation case (545 diseases without validation cases are not shown), coloured by the step at which each disease left the selection path. Dashed lines show the AUROC and sensitivity cutoffs. **c**, Detection rates among all first-diagnosis cases for the 33 diseases with sufficient test support (28,828 held-out participants; 62,820 participant–disease episodes), in the order of Fig. 2i; *n* gives cases per disease. Perineal laceration has too few high-history-burden cases to appear in Fig. 2i. **d**, Detection rates in high-history-burden versus all cases for the 32 diseases at 120 and 10 days before diagnosis; the dashed line denotes identity. **e**, Median disease-level detection rate among all diagnosis-aligned cases (left) and median positive predictive value (PPV) at fixed test visits (right) at three validation-specificity cutoffs. Detection rates in a–d use the 95% cutoff. The two parts of e use different query populations and cannot be combined into a single confusion matrix; these retrospective sensitivities are not positive predictive values.

**Figure S7:**
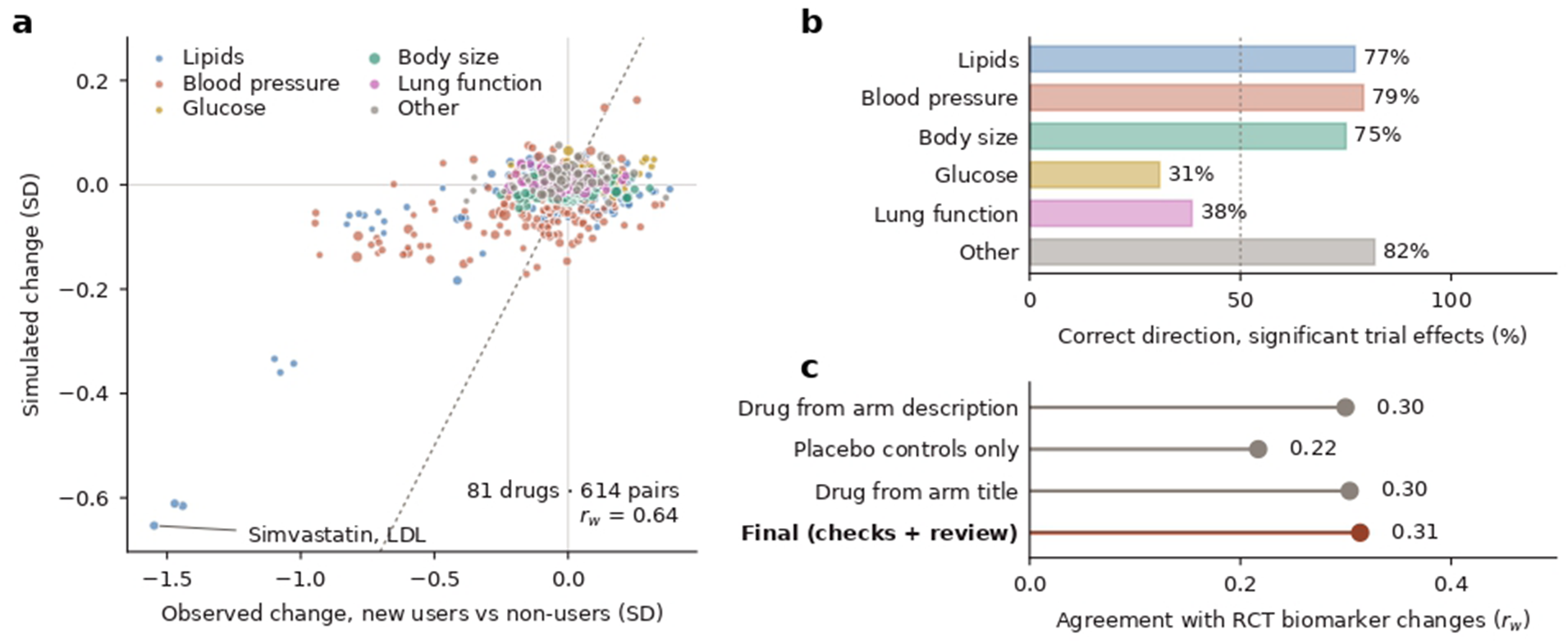
Simulated physiological responses to medications. **a**, Simulated versus observed biomarker change in held-out UK Biobank participants who started each of 81 drugs, relative to non-users over the same interval (614 drug–biomarker pairs). Point size reflects precision and colours denote biomarker class; the dashed line denotes identity. **b**, Proportion of significant trial effects (*P* < 0.05) among the drug–biomarker pairs in Fig. 4b whose direction was reproduced by the simulation, by biomarker class. The dashed line marks 50%. **c**, Agreement with RCT biomarker changes under alternative rules for extracting trial data; the final rule is used in Fig. 4b. *r_w_*, precision-weighted correlation; SD, UK Biobank standard deviation.

**Figure S8:**
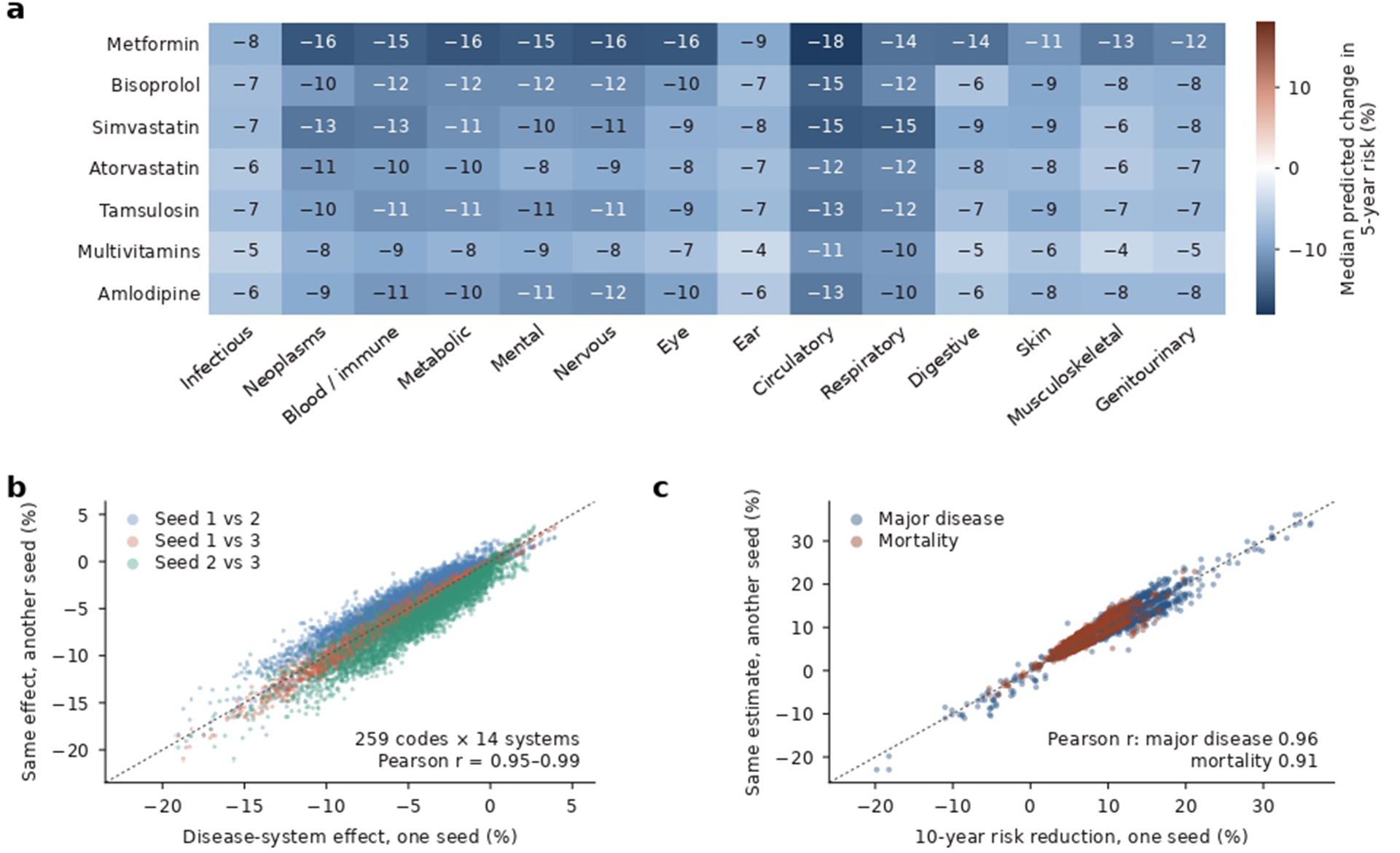
Disease-system profiles and reproducibility of the medication screen. **a**, Median predicted change in 5-year risk across 14 disease systems for the candidate medications in Table 2, simulated in their held-out new users. **b**, Disease-system effects for all 259 medication codes estimated by independently trained models (three random seeds), compared pairwise. **c**, Ten-year major-disease and mortality risk reductions in the shared reference cohort, compared pairwise between seeds. Dashed lines denote identity.

